# Individual vaccine efficacy variation with time since mRNA BNT162b2 vaccination estimated by rapid, quantitative antibody measurements from a finger-prick sample

**DOI:** 10.1101/2021.11.30.21267102

**Authors:** Matheus J. T. Vargas, Mithileshwari Chandrasekhar, Yong Je Kwon, Gerrit Sjoerd Deijs, Carsten Ma On Wong Corazza, Angela (Wai Yin) Chai, Rebecca L. Binedell, Ellen Jose, Bhavesh Govind, Laura Huyet, Pooja K. Patel, Gabrielle Reshef, Vijaya Kumar, Tiffany Lowe, Robert J. Powell, Kieran C. Jina, Flynn C.W. Walker, Apisalome Talemaitoga, M. Cather Simpson, David E. Williams

## Abstract

We show that an individual’s immune status to Covid-19 can be monitored through quantitative antibody measurements using a method based on centrifugal microfluidics, specifically designed for speed to result (20 min), high throughput (8 samples simultaneously) and accuracy from a finger-prick blood sample. Anti-Receptor Binding Domain (RBD) IgG concentration showed a log-normal distribution with mean decreasing with time following the second vaccination with mRNA BNT162b2 (Pfizer). Using a model for an individual’s antibody concentration-dependent vaccine efficacy allowed comparison with literature data on changing vaccine efficacy against symptomatic disease across a population. Even though the trial was small (*n* = 100) the computed population vaccine efficacy was in reasonable agreement with that obtained from a large population survey. The derived parameters for the vaccine efficacy model were in good agreement with those expected from previous studies and from a simple theoretical model. The results and modelling show that the major proportion of breakthrough infections are for people whose antibody concentration is in the tail of the distribution. The results provide strong support for personalized booster programmes that, by targeting people in the tail of the distribution, should be more effective at diminishing breakthrough infection and optimising booster dose supply than a program that simply mandates a booster at a specific post-vaccination time point.

## Introduction

The Orbis high-throughput quantitative immunity measurement system implements an enzyme-linked immunosorbent assay adapted for accurate, precise, rapid-testing (<20 min) on a centrifugal microfluidic platform. All operations post sample collection and loading onto the instrument are automated. The sample is a small drop of whole blood obtained from a finger-prick. The assay is designed for speed – multiple samples are measured at the same time with a total assay time less than 20 min – and for accuracy. The important elements of the assay design are rapid, complete mixing and accurate timing. These features mean that a kinetically-controlled assay can be implemented: concentration is deduced from the rate of binding (‘on’ rate) of the target to the capture surface; a wide, linear dynamic range is achieved. The capture surface is a single small bead that carries the SARS-CoV-2 – Receptor Binding Domain (RBD) protein so the measurement is of IgG concentration directed at this key protein. Speed of mixing is obtained by rapid oscillation of the disc motion to cause oscillation of the bead within the chamber, whose shape is also controlled to promote mixing *(1)*. The assay performs with high analytical sensitivity, therefore samples are diluted in the machine with 10x addition of buffer. The buffer dilution step also minimises effects of non-specific adsorption of blood components. The details of the assay design and results on buffer solutions are given in the Supporting Information, SI, Supplementary Appendix 2, demonstrating repeatability of duplicate measurements of 10% across a range of 5 – 500 ng/mL of IgG.

As part of the device development, a small clinical trial was conducted *(2)*. The primary outcome was to demonstrate the system’s ability to distinguish fully vaccinated from unvaccinated people by means of a simple and rapid procedure, that might for example be used for entry control or validation of a vaccine passport: an approach of intense current interest. Vaccinated participants had a range of age and time since completion of vaccination (2 doses separated by at least 3 weeks). Because of the nature of the primary outcome, there was no selection by age, date since vaccination, ethnicity or socio-economic factors. Participant age and time since vaccination were recorded as was the experience of the participants and sampling system operators during the process of taking the sample. The resulting data provided a distribution across the sample of anti-SARS-CoV-2 – RBD IgG concentration. This paper presents the results of an analysis of the change in antibody concentration distribution with time since vaccination.

## Results

The output of the measurement is the concentration of anti-RBD IgG with respect to a buffer control that is measured simultaneously. Assay validation used buffer controls and the panel of samples provided by the National Institute of Biological Standards and Controls, UK, NIBSC 20/B770. This panel contains as a package insert the results of analyses using a number of different assay platforms. The Orbis assay result was converted to Binding Antibody Units (BAU) for the RBD using WHO and NIBSC standards NIBSC 20/162, 20/150, 20/148 20/144 and 20/140. Correlation of Orbis assay results with the standards, comparison of results for different assay platforms using the data provided with NIBS 20/B770, and comparison of Orbis assay results with the data provided with NIBS 20/B770 is given in Supplementary Appendix 3. These results show that the Orbis assay gives results that are comparable with commercial assay platforms.

The distribution of anti-RBD IgG concentrations for vaccinated and unvaccinated participants is shown in Supplementary Fig 1. The results for vaccinated participants showed a log-normal distribution of concentration. Unvaccinated participants also showed a signal, but this was significantly smaller and normally distributed. The results for the NIBSC standards showed the same effect, with the same mean offset and offset standard deviation. Antigen (RBD) titration and measurement using buffers containing fibrinogen showed that this result was due to non-specific adsorption of the secondary indicator antibody promoted by fibrinogen adsorption to the assay chamber walls in the version of the assay used for the trial, where prevention of non-specific binding had not been optimised. In the following, the mean value from this normally-distributed background effect has been subtracted from the assay results for vaccinated participants, and the distribution of the non-specific background results has been incorporated in the error model. The assay also shows a replication error (difference between duplicate measurements) that is constant at 10% across the assay range: supplementary figure 1C

Participant age was fairly uniformly distributed across the range 20 – 60 yr. The vaccination date correlated data fall into 3 clearly distinguishable ranges (fig 1 inset): < 90 days, 90 – 170 days and 170 – 228 days. Figure 1A shows the empirical cumulative probability distribution of anti-RBD IgG concentration for these 3 ranges. Figure 1A also shows the fit to a log-normal distribution. The log-normal mean decreases with time since vaccination. The effect is highly significant (<0.5% for the Kolmogorov-Smirnov test) between < 90 days and 90 – 170 days, and marginally significant (10% K-S) between 90 – 170 days and 170 – 228 days. Bootstrap sampling estimation of the ln-normal mean clearly showed the decrease with time : Figure 1C. The ln-normal standard deviation was constant with time: *σ* = 0.59 ±0.02. An effect of age correlated with time since vaccination was marginally discernable in the data but without statistical significance. Thus the following discussion focusses on the effect of time since vaccination.

**Figure 1.**
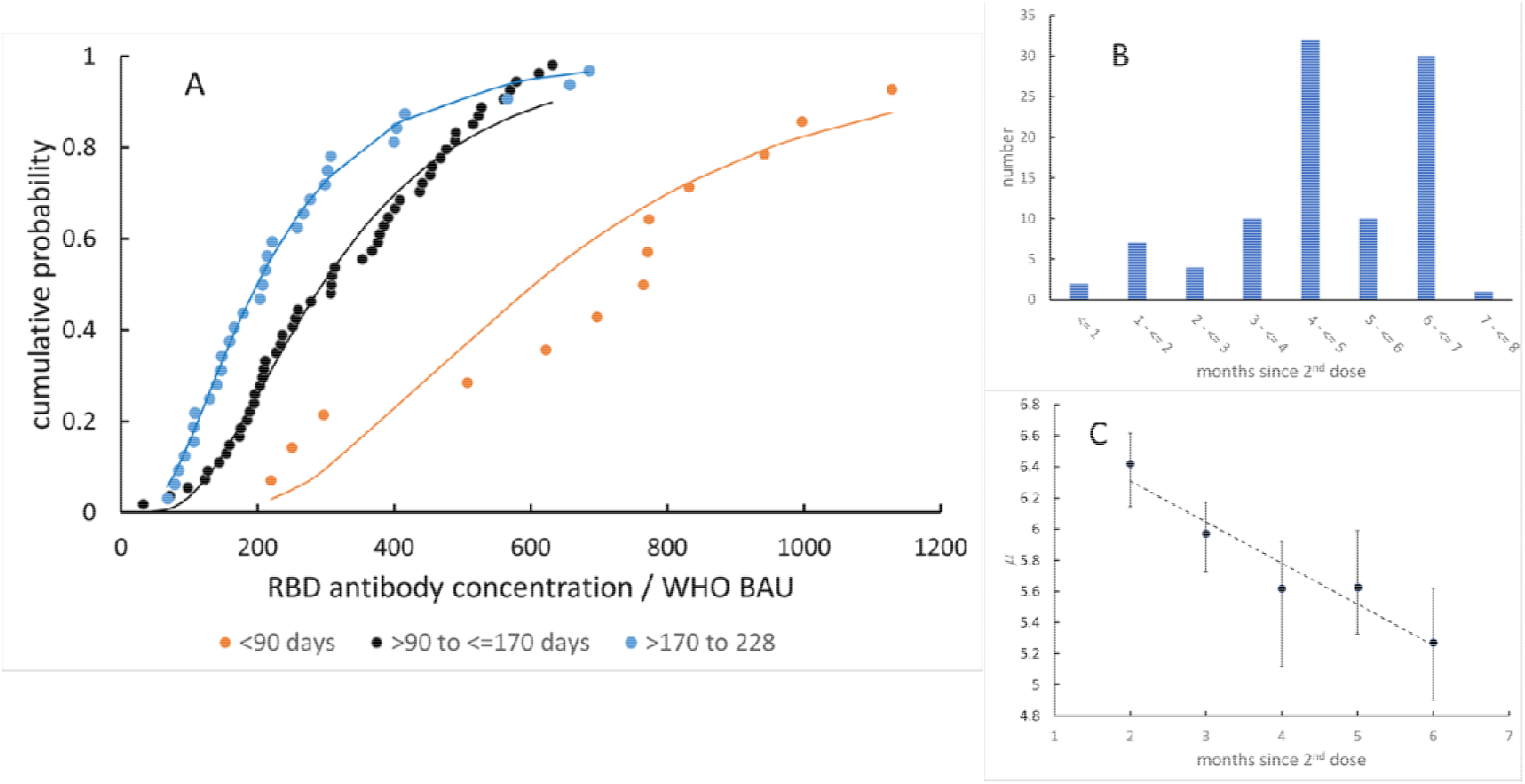
A: Anti-RBD IgG concentration distribution (offset subtracted), and its alteration with time since vaccination. The lines are fits to a ln-normal distribution. B: distribution of vaccination dates (2^nd^ dose) by month across the study participants. C: variation of mean of ln-normal concentration distribution (concentration in BAU) with time since 2^nd^ dose (data labelled 6 months are all data > 5 months), estimated by bootstrap sampling.

## Discussion

Vaccine efficacy against symptomatic illness is now clearly known to decay with time following completion of vaccination, significantly over the first 6 months *(3, 4)*. The large study by Tartof et al.*(3)* provides good information about the breakthrough infection probability for a population, expressed as vaccine efficacy, VE, as a function of time following vaccination, for variants through to Delta. Given the antibody concentration distribution shown in Figure 1, assuming that the probability of symptomatic infection is determined by the binding of IgG to the RBD *(5)* and using a model relating symptomatic infection probability to the anti-RBD concentration *(5, 6)*, the population breakthrough infection probability and hence population VE can be calculated from the data collected in the study reported here. Estimation of the model parameters to match the calculated population VE with that observed by Tartof et al. allows calculation of an estimate of individual VE given the individual anti-RBD concentration. The detail of the calculation including the bootstrap sampling method used to estimate the confidence intervals is given in Supplementary Appendix 1. Briefly, the model *(5, 6)* is:
(1)
where *E*_*I*_*(c)* denotes individual vaccine protective efficacy as a function of concentration, *c*, of anti RBD IgG. The curve is a sigmoidal variation of ln*(c)*. The parameters are *c*_*50*_ and k where *c*_*50*_ denotes the concentration for 50% vaccine efficacy and *k* controls the rate of increase of efficacy with concentration around *c*_*50*_. The population VE is calculated by integration over the population distribution of antibody concentration. The aim is to find a set of parameters that is not only consistent with the population VE for different date ranges, but also consistent with the observed antibody concentration distribution across the different date ranges and with model constraints on *c*_*50*_ and *k (5-7)*. Figure 2 shows the results and Table 1 lists the parameters and compares the observed and calculated population VE. The estimate of k is not dependent on the antibody concentration units used. Khouri et al*(6)* give *k* = 1.30 with 95% confidence interval 0.96 – 1.82. Williams(5) has shown that this range is consistent with a simple physical model for antibody protection. The estimated k is very consistent with this value. Determination of *c50* is dependent on the concentration scale used. In order to avoid this difficulty, Khouri et al gave the values as a multiple of the median convalescent antibody concentration, *c*_*50*_ = 0.2 (0.14,0.28). The convalescent median assessed from the 23 samples in the NIBSC 20/B770 panel by the Orbis device is 370 BAU. The Khouri et al. result would thus give *c*_*50*_ =74 (51, 103). The estimated *c*_*50*_ is higher. The Khouri et al. result was estimated for the original (Wuhan) variant. The data used here from Tartof et al *(3)* are an average for all variants present in California up to and including Delta. The value for c_50_ would increase with decreasing antibody affinity for the spike protein *(5)*. Wall et al.*(7)* have suggested qualitatively that *c*_*50*_ for the Delta variant could be a factor of 6 times higher, implying c_50_ ∼ 440 BAU. The estimated *c*_*50*_ is indeed consistent with this number, given the range of variants present in the study population of Tartof et al.

**Table 1.** Observed and calculated population vaccine efficacy. The parameters for the (natural) log-normal concentration distributions are μ and σ where the concentration is expressed in BAU. The derived parameters c_50_ and k are consistent with previous studies(5-7).

| Time range /month | $\leq 2$ | $2 - \leq 3$ | $3 - \leq 4$ | $4 - \leq 5$ | $> 5$ |
| --- | --- | --- | --- | --- | --- |
| $\mu$ | 6.4<br>(6.1,6.6) | 6.0<br>(5.7,6.2) | 5.6<br>(5.1,5.9) | 5.6<br>(5.3,6.0) | 5.3<br>(4.9,5.6) |
| $\sigma$ | 0.59 ± 0.02 | | | | |
| Population<br>VE(observed: Tartof)/<br>% | 83<br>(80,86) | 77<br>(76,78) | 68 (66,70) | 61<br>(59,64) | 47<br>(43,50) |
| Population<br>VE(calculated) | 79<br>(68,89) | 70<br>(60,82) | 57 (40,70) | 58<br>(45,69) | 47<br>(34,62) |
| $c_{50}$ / BAU | 212 (151,272) | | | | |
| $k$ | 1.56 (1.11,2.02) | | | | |

**Figure 2.**
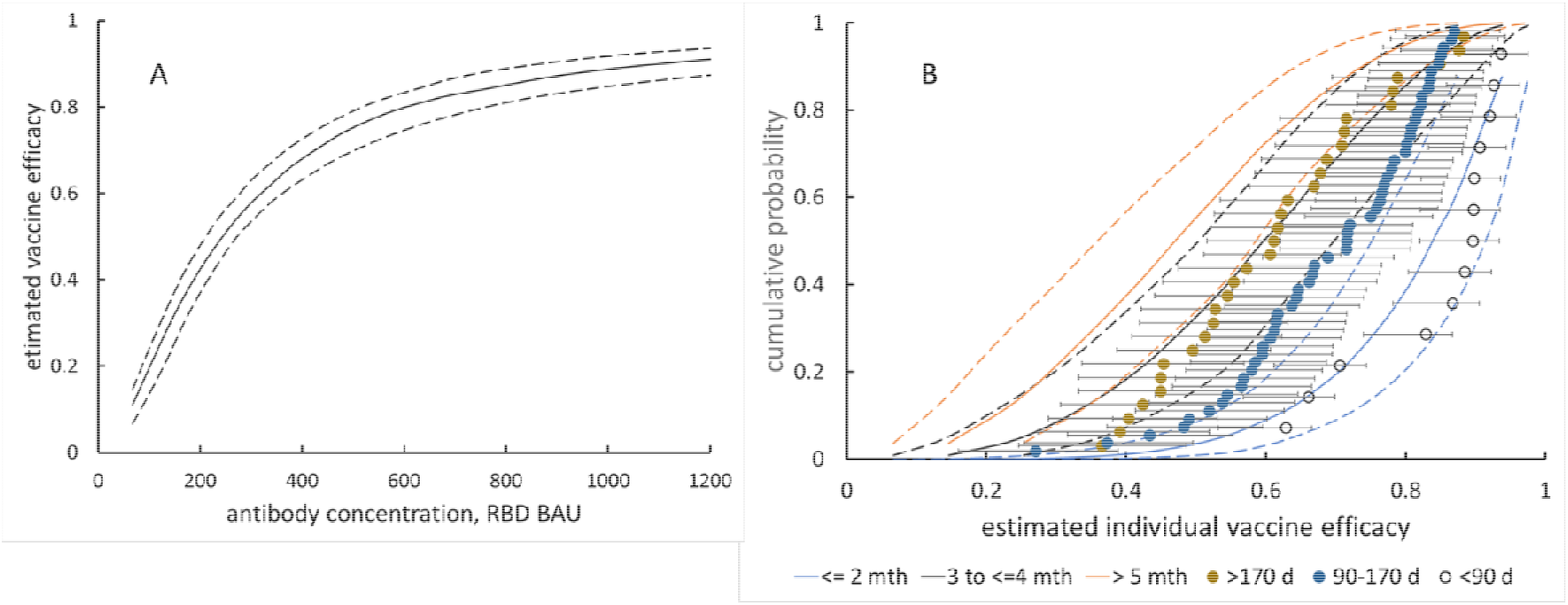
A: Estimated vaccine efficacy against antibody concentration, derived by fitting of equation (1) ; dashed lines give 95% confidence intervals. B: Empirical cumulative probability distribution of vaccine efficacy across different ranges of time since vaccination, estimated using the data of Tartof et al (3), the concentration distributions shown in Figure 1 and the calculation given in Supplementary Appendix 1. The lines are the fitted theoretical distributions; dashed lines and error bars give 95% confidence intervals.

The limitation of this study is that the small number of participants, without comprehensive coverage of the full range of vaccination date, limited the accuracy of assessment of the probability distribution of concentration for the full range of date. However, even with this small number of participants the model fit is reasonable and shows how observations of breakthrough infections combined with quantitative measurement of antibody concentration and the simple model employed here can be used to assess individual vaccine efficacy and hence individual risk in the face of the continuing development of the pandemic and emergence of new variants. Since the value of k is reasonably constrained, this model asserts that assessment of vaccine efficacy against new variants comes down to an assessment of antibody binding affinity to the RBD, which would then lead to a revised value for the parameter *c*_*50*_.

Figure 2 illustrates the very significant range of estimated VE across the participants in this study. The effect of the log-normal distribution of concentration is that the most significant contribution to breakthrough infections comes from people in the low-concentration tail of the distribution. The results have implications for the design of booster vaccination programmes directed at optimising the use of booster doses in order to achieve the best possible population protection. Since the most significant contribution to breakthrough infections comes from people in the low-concentration tail of the distribution, it is arguable that any booster programme should be directed at these. It would arguably be less useful to boost those in the high concentration tail. Significant numbers of people can be found in the low-concentration tail relatively recently after completion of vaccination whilst others can be found in the high concentration tail even after 6 months. Therefore a vaccination programme that simply targets all people > 6 months after vaccination, whilst administratively convenient, will miss an important proportion of the population at risk and will also unnecessarily boost others. There is now a focus on quantitative antibody tests that promote an increased understanding of immune responses to SARS-CoV-2 *(8)*. The work described here has demonstrated a fast and accurate, point-of-need, finger-prick immunoassay method that overcomes the well-known limitations of lateral-flow rapid assay methods and is appropriate for large-scale studies. Despite the limitations imposed by the small sample size, particularly the lack of resolution in date range over a full 6 months leading to uncertainty in model parameter estimation, the work has demonstrated that a rapid antibody measurement can build the evidence needed to construct the targeted booster programmes that are now being called for *(9)*.

## Supporting information

spllementary figures and appendices

## Data Availability

All data produced in the present work are contained in the manuscript or supplementary appendices

## Acknowledgement

DEW acknowledges early support through an E.T.S Walton Visiting Fellowship of Science Foundation Ireland. The work has been funded by Orbis Diagnostics. We thank the staff of Cavendish Clinic, Manukau, Auckland for their management, sample collection and interaction with the participants for the clinical trial, and particularly thank the participants themselves. The prototype instrument used for this trial was designed and constructed by D&K Engineering, San Diego, California; particular thanks to David Kortbawi, Sean McCrossin, Victor Escobedo, Don Fox and John Muren.

