## Supplementary material for "Individual vaccine efficacy variation with time since mRNA BNT162b2 vaccination estimated by rapid, quantitative antibody measurements from a finger-prick sample": spllementary figures and appendices

**This PDF file includes:**

Supplementary Figures S1

Supplementary Appendix 1: Estimating individual vaccine efficacy from population breakthrough risk

Supplementary Appendix 2: Assay design

Supplementary Appendix 3: Assay validation with WHO standards

Supplementary Figures

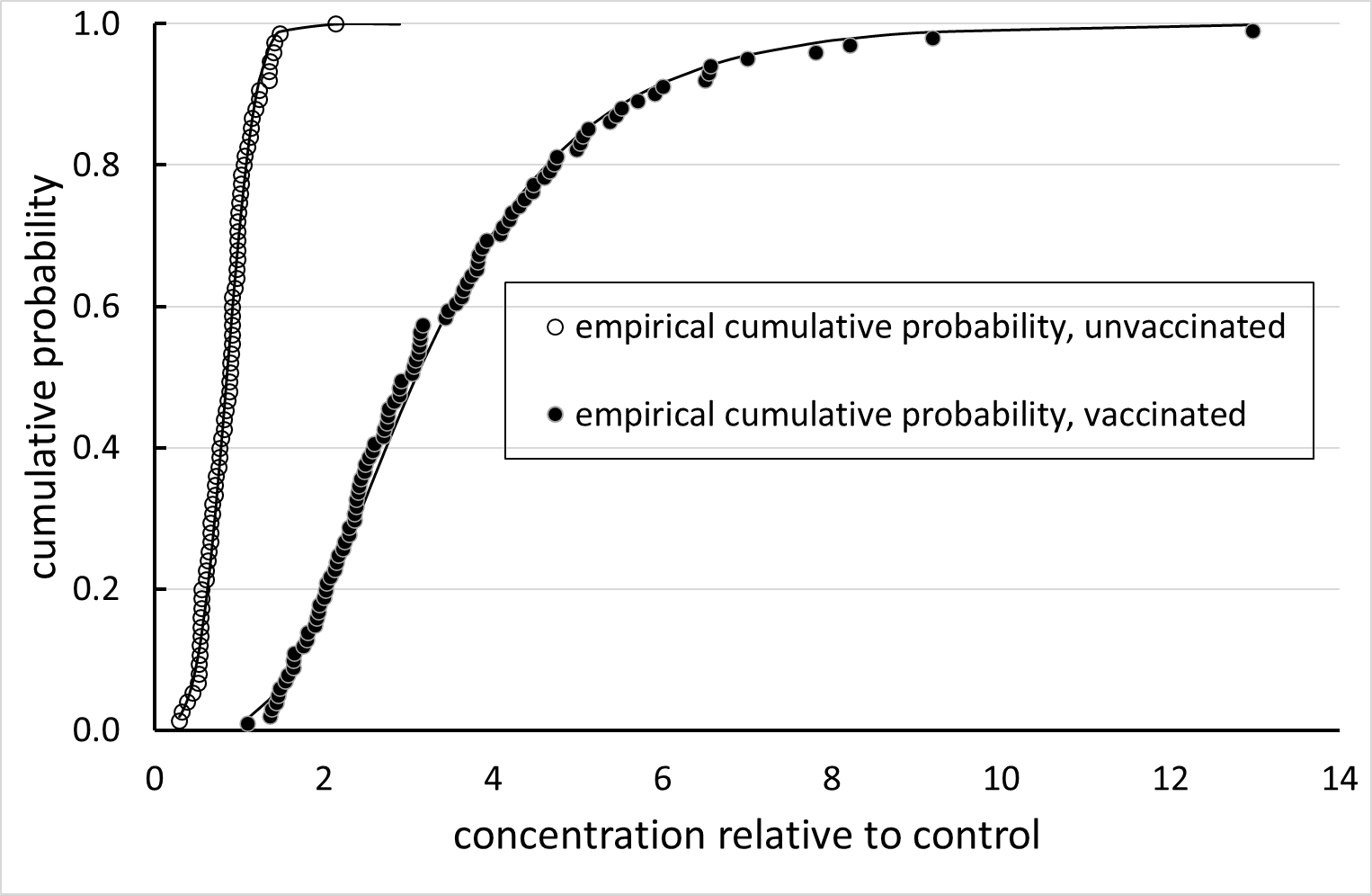

Supplementary Figure 1A: Raw data distributions comparing Orbis device signal for unvaccinated with that for vaccinated study participants. The lines show the fit to a normal distribution for the unvaccinated and a log-normal distribution for the vaccinated results.

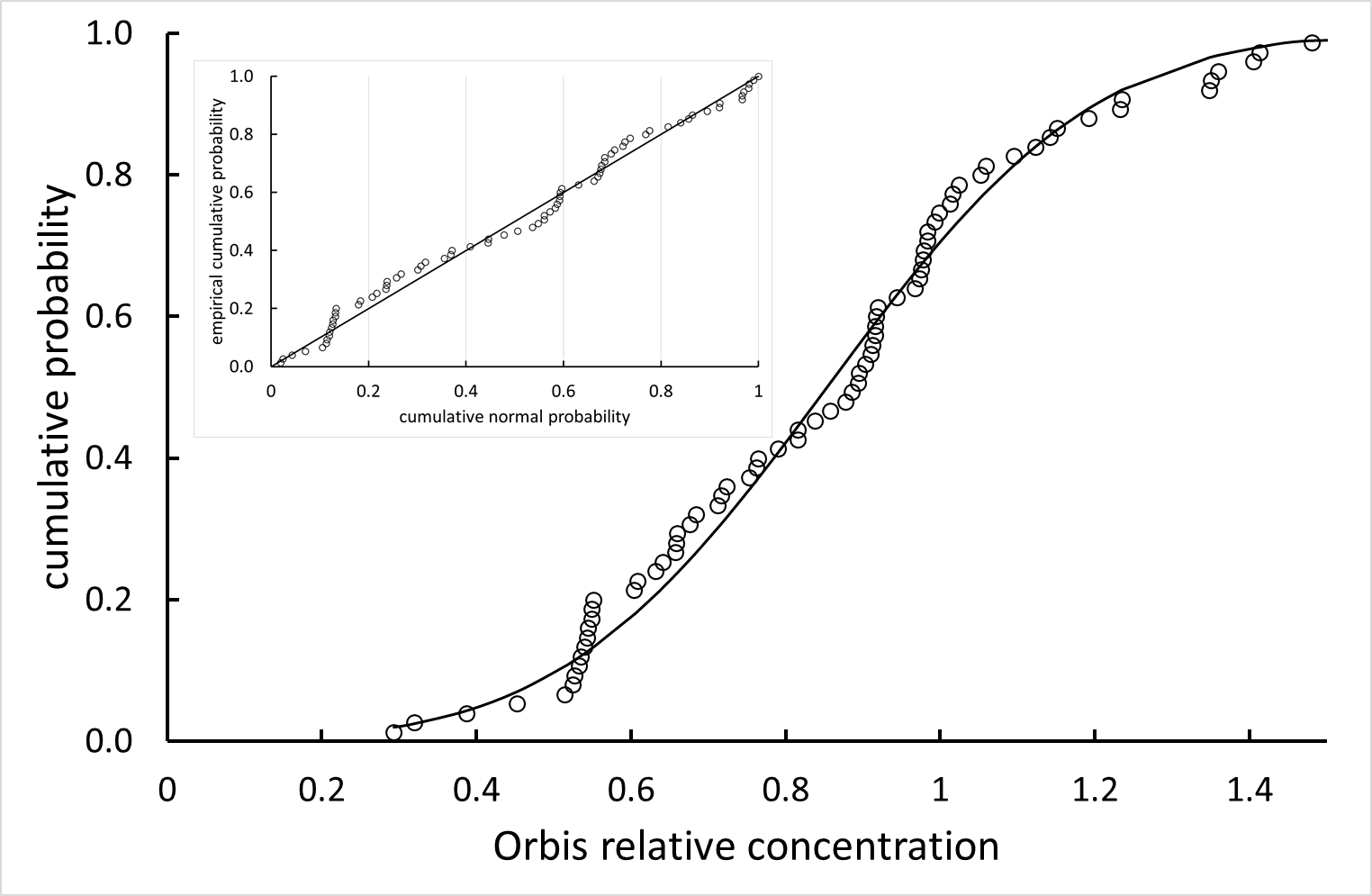

Supplementary Figure 1B: Comparison of Orbis device signal for unvaccinated participants with that for a normal distribution (mean = 0.85 standard deviation = 0.27 in relative concentration units)

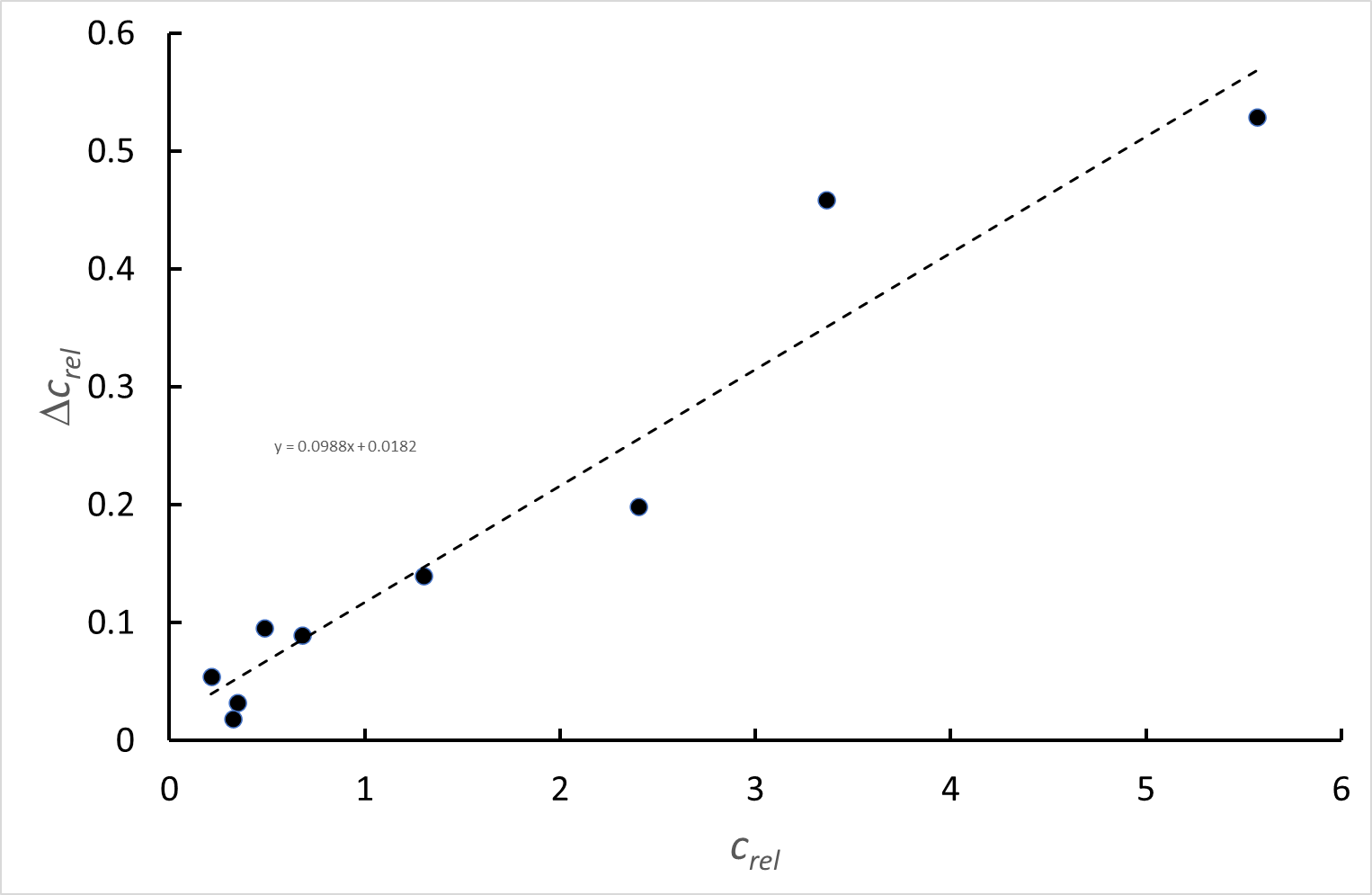

Supplementary Figure 1C Difference between duplicate measurements relative to the control, Δ*c_rel_* as a function of the measured concentration relative to the control, *c_rel_* ; the assay shows a coefficient of variation of 10% that is essentially constant across the assay range

**Supplementary Appendix 1: Estimating individual vaccine efficacy from population breakthrough risk**

1. Vaccine efficacy model for an individual

Khouri et al.(*1*) have described empirically the dependence of vaccine protective efficacy on neutralising antibody concentration. Williams has shown how the observed dependence has a simple physical explanation in the statistics of binding of antibody to the viral spikes (*2*). Cromer and others have used this model to describe the changes in vaccine efficacy with different variants (*3, 4*) : according to the ideas put forward by Williams (*2*), the effect is simply due to change in binding affinity, and binding affinity distribution, of the antibodies to the viral spike . Therefore it seems reasonable to use this model to relate the concentration of anti-receptor binding domain (RBD) IgG to vaccine efficacy, perhaps with parameters slightly different to those derived by Khouri but which should be consistent with the model developed by Williams.

The model is:

$$E_{I}\left( c \right)=1/\left[ 1+\left( {c_{50}}/c \right)^{k} \right]$$

(1)

Where *E_I_(c)* denotes vaccine protective efficacy as a function of concentration, *c*, of anti RBD IgG. The curve is a sigmoidal variation of ln(*c*). The parameters are *c_50_* and *k* where *c_50_* denotes the concentration for 50% vaccine efficacy and *k* controls the rate of increase of efficacy with concentration around *c_50_* . In what follows, *c_50_* and *k* will be adjusted to match observed results for population breakthrough infection.

2. Breakthrough infection risk by integrating across population antibody concentration distribution

Williams (*5*) used equation (1) to derive population breakthrough risk given a known antibody concentration distribution across the population.

$$P\left( protected \right)=\int_{0}^{\infty} E_{I}\left( c \right)f_{V}\left( c \right)dc$$

(2)

Where *f_V_(c)* denotes the probability density of concentration, *c*, across the population. As demonstrated, this distribution is log-normal, consistent with the observations of Khouri et al.

$$f_{V}\left( c \right)\mathbb{\sim N}\left( ln(c) | \mu,\sigma\right)$$

(3)

Where the mean, *μ*, and standard deviation, *σ* , may vary with time since vaccination.

Therefore, if there are sufficient data to estimate the mean, *μ*, and standard deviation, *σ* , over a range of time since vaccination, then an estimate of vaccine efficacy, *VE*, over this time range can be obtained:

$$VE=P\left( protected \right)=\int_{0}^{\infty} \left( 1/\left[ 1+\left( {c_{50}}/c \right)^{k} \right] \right)\mathbb{N}\left( ln(c) | \mu,\sigma\right)dc$$

(4)

If the vaccine efficacy over this time range is known from population infection statistics comparing vaccinated and unvaccinated people, then if data for enough time ranges is available the parameters *c_50_* and *k* may be adjusted to obtain agreement between observed and computed efficacy. The functions and integral can be evaluated in a Microsoft Excel spreadsheet. The unknown parameters *c_50_* and *k* can be estimated by optimising the match between observed and calculated VE for the different time ranges for which data are available, with adjustment of the parameters *μ* and *σ* to obtain consistency between the distribution of vaccine efficacy calculated for the samples from equation (1) and the assessment of distribution of antibody concentration derived from fitting the measurements for the samples, represented by equation (3).

3. Use of distributions over different date ranges to derive parameters of vaccine efficacy model, using vaccine efficacy data over time since vaccination, from Tartof et al. (*6*)

Tartof et al obtained vaccine efficacy against infection for the Pfizer-BioNtech vaccine varying with time since vaccination using data scoured from medical records of over 3M people. Results averaged across all variants are given in Table 1

Table 1

| Vaccine efficacy against symptomatic infection, un-adjusted for age. ethnicity and other factors(*6*) | | | | | |
| --- | --- | --- | --- | --- | --- |
| < 1 month | 1 to <2 | 2 to <3 | 3 to <4 | 4 to <5 | >=5 |
| 88 (86-89) | 83 (81-85) | 77 (76-79) | 68 (65-70) | 61 (58-64) | 47 (43-51) |

Figure 1 shows the distribution of time since second dose across the clinical study participants. Within each time range, where the minimum was taken as ≤2 months and the maximum as > 5 months, the offset-subtracted data were sampled by the bootstrap method with replacement to generate 200 independent samples for which the log-normal mean and standard deviation of anti-RBD IgG concentration was calculated. For each cycle of sampling, a Gaussian random error was added, with two components: a relative error to reflect the replication error observed in the buffer control studies, and an error in the offset to reflect the variability of the non-specific binding component of the signal, as determined from the samples for un-vaccinated participants. Figure 2 shows the variation with date since second dose of the bootstrap natural log mean and standard deviation. The mean decreased monotonically with time since vaccination. The standard deviation was constant though with significant estimation error. Each cycle of calculation of vaccine efficacy took one bootstrap sample of natural log mean and standard deviation for each time range, together with a bootstrap sample from the data of Tartof et al for each time range. Given these values, the integral of equation (4) was evaluated for each time range and the parameters *c_50_* and *k* adjusted to minimise the squared deviations of calculated and observed VE across the five time ranges, weighting the squared deviation in each time range with the number of participants falling into each time range. This procedure generated 200 bootstrap estimates of the vaccine efficacy for each time range and of the parameters *c_50_* and *k* . Figures 3 and 4 show the results.

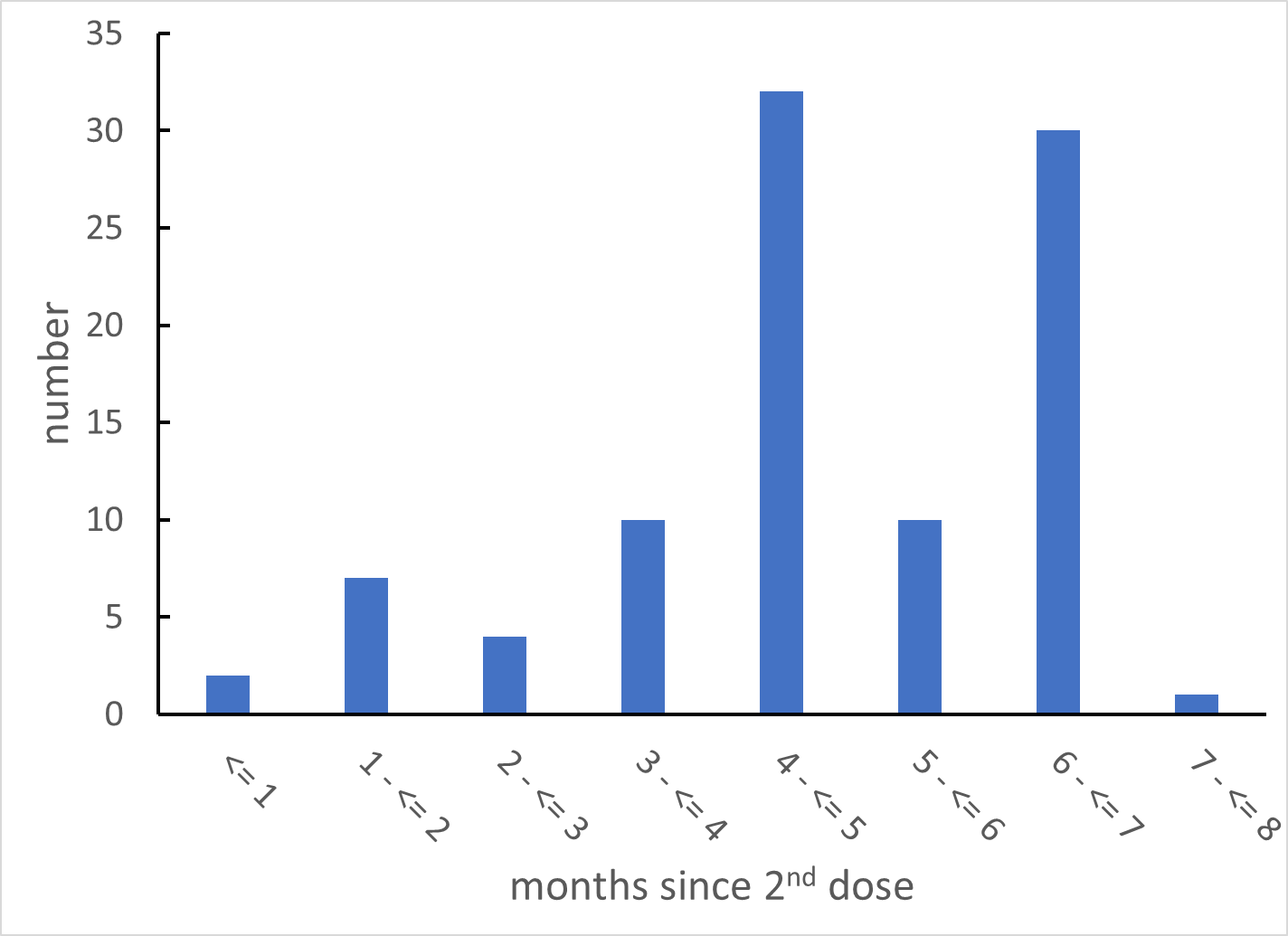

Figure 1: Distribution of number of study participants across months since second vaccine dose.

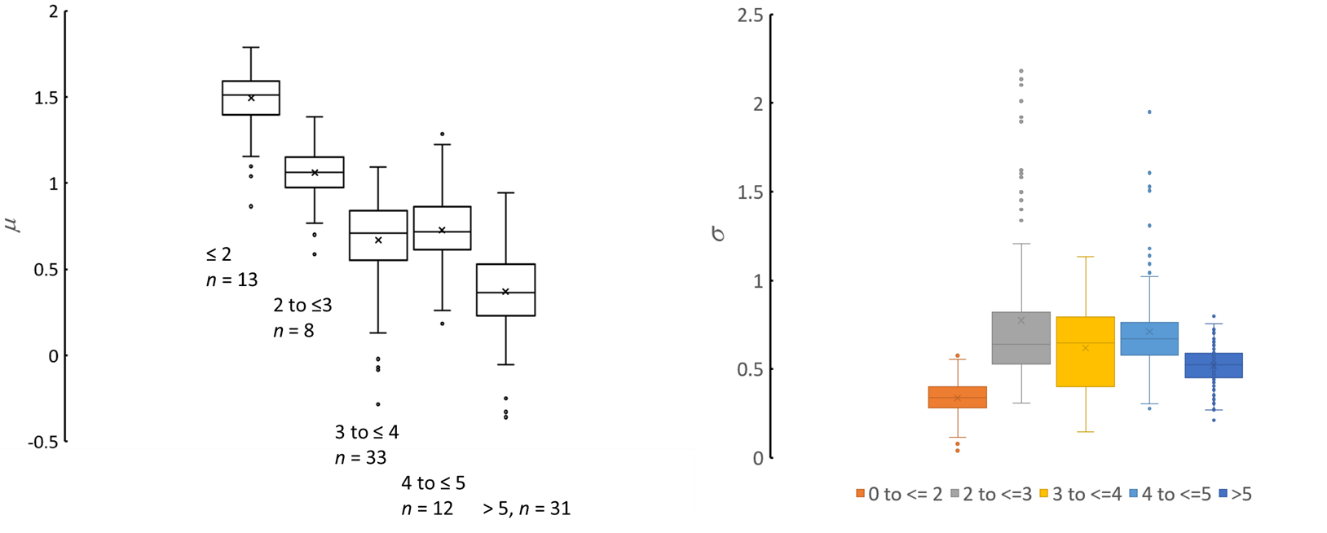

Figure 2. Bootstrap sample natural log mean, *μ* (orbis units : 1 unit = 136 RBD BAU) and standard deviation, *σ* , for different times since 2^nd^ vaccination.

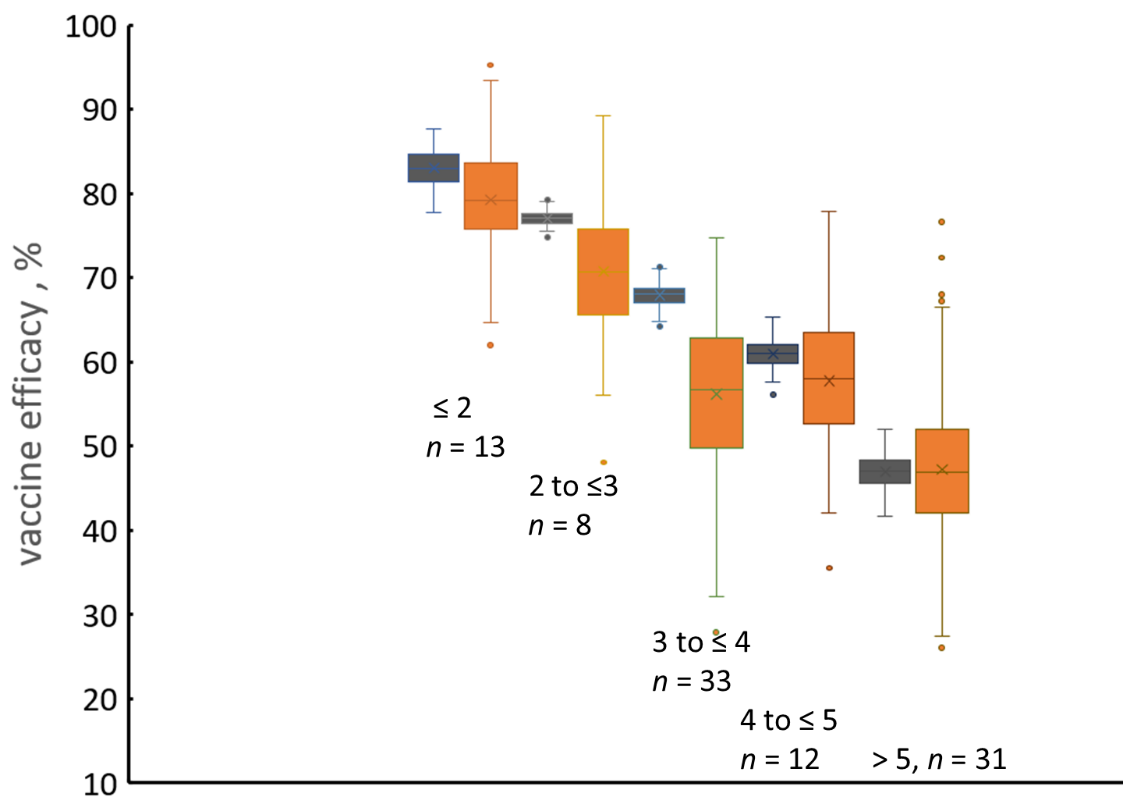

Figure 3. Vaccine efficacy for the different time ranges since vaccination (months). Black: bootstrap samples taken from the data of Tartof et al (*6*); Orange : calculated from bootstrap samples taken from the present data on antibody concentration distribution variation with time, fitting the parameters *c_50_* and *k*.

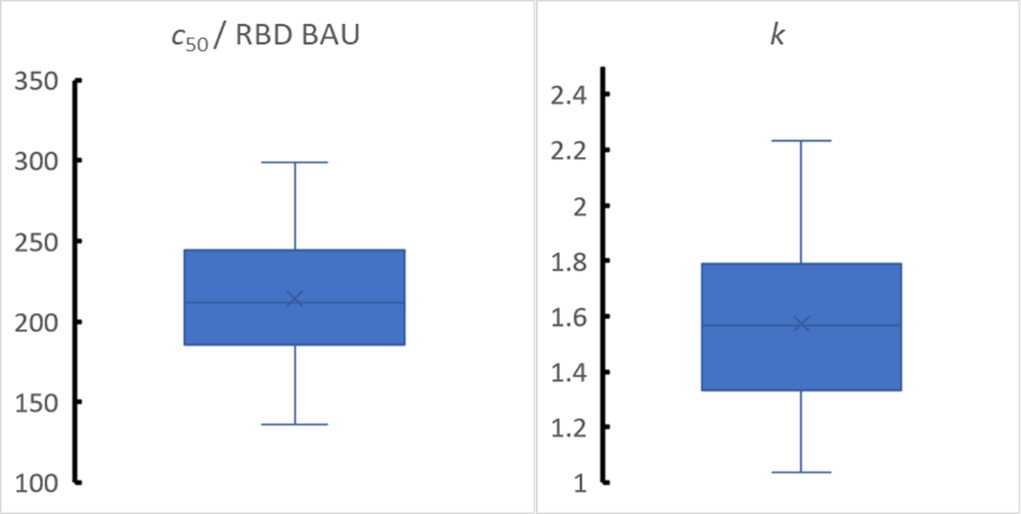

Figure 4. Distribution of values of *c_50_* and *k* computed using the bootstrap procedure

4. Reasonableness of parameters.

The estimate of *k* is not dependent on the antibody concentration units used. Khouri et al (*1*)give *k* = 1.30 with 95% confidence interval 0.96 – 1.82. Williams (*2*) has shown that this range is consistent with a simple physical model for antibody protection. The estimated *k* (95% confidence interval) is 1.56 (1.11, 2.01) is very consistent with this value. Determination of *c_50_* is dependent on the concentration scale used. In order to avoid this difficulty, Khouri et al gave the values as a multiple of the median convalescent antibody concentration, *c_50_* = 0.2 (0.14,0.28). The convalescent median assessed from the 23 samples in the NIBSC 20/B770 panel by the Orbis device is 370 BAU. The Khouri et al. result would thus give *c_50_ =*74 (51, 103) . The estimated *c_50_* is higher: 212 (151,273). The Khouri et al. result was estimated for the original (Wuhan) variant. The data used here from Tartof et al are an average for all variants present in California up to and including Delta. The value for *c_50_* would increase with decreasing antibody affinity for the spike protein (*2*). Wall et al.(*7*) have suggested qualitatively that *c_50_* for the Delta variant could be a factor of 6 times higher, implying *c_50_* ~ 440 BAU. The estimated *c_50_* is indeed consistent with this number, given the range of variants present in the study population of Tartof et al.

Supplementary Appendix 2: Assay design.

1. *Assay principles*

Immunoassay of the ‘sandwich’ or ‘indirect ELISA’ type comprises the following steps:

(a) Primary binding step - Mixing of the sample with a surface that has binding sites specific for the target analyte (eg a surface carrying bound antigen to a target antibody). The target binds to sites on the surface. After some time, the reaction is stopped and any unbound material is washed away.
(b) Incubation of the surface carrying the captured target with a label that binds only onto captured target. After some time the reaction is stopped and any unbound material is washed away. The amount of label remaining on the surface is measured, thus giving the amount of captured target and hence the original sample concentration.

There are two contrasting methods for the primary binding step, for connecting the solution concentration to the amount of captured analyte:

(a) The primary binding step is run for a time sufficiently long that the surface reaches equilibrium with the solution (titration method)
(b) The primary binding step is run for a short time: the amount captured is determined by the kinetics of reaction of the analyte with the surface. For an accurate determination, this method requires precise control of mixing, which should be very rapid in comparison with the time scale of the reaction (determines time zero and concentration uniformity during the reaction) and precise control of reaction time.

The determining time scale for an antibody-antigen reaction is the ‘off’ rate: the rate of dissociation of antibody from antigen (typically 10^-3^ s^-1^) . For the equilibrium titration method, the time for the primary binding step needs to be several times the reciprocal of the ‘off’ rate: so at least 1 hr. For the kinetic method, the time scale for the primary binding step can be short in comparison with the reciprocal of the ‘off’ rate. Because the surface-bound antibody-antigen complex can dissociate during the washing and secondary binding steps, the time scale for these steps should be short in comparison with the reciprocal of the ‘off’ rate, and well-controlled.

The Orbis assay is a kinetic assay. The key is rapid and complete mixing upon addition of both the sample and the label to the capture surface, and precise timing of each step. The following sections develop the theoretical model for this type of assay.

2. *Kinetic assay model*

The capture surface has a total number, *N_A_*, of capture sites available. These sites are assumed sufficiently widely spaced that they are independent of one another – there is no interaction between sites that alters the probability of reaction of a given site dependent on the occupation of neighbouring sites. At any time ,*t*, following the start of the reaction, a fraction, *θ*, of these sites are occupied by analyte. Hence the number of empty sites at time, *t*, is (1-*θ* )*N_A_* . The analyte has solution concentration, *c*, with value *c_0_* at the start of the reaction (*t* = 0); *c_0_* is the quantity to be determined by the assay procedure. The rate constant for attachment of analyte is *k_on_* and for detachment is *k_off_* .

The treatment below assumes that the number of binding sites is sufficiently large that there is no need to develop a stochastic model (that is, to count the binding events as discrete steps, one molecule at a time). A continuum model is given.

Primary binding step: For the primary binding step:

$$\frac{d\theta}{dt}=k_{on}c\left( 1-\theta\right)-k_{off}\theta$$

(1)

The total amount of analyte, bound and free, is fixed, so:

$$c_{0}=c+\frac{\theta N_{A}}{V}$$

(2)

$$\frac{d\theta}{dt}=\left[ k_{on}\left( c_{0}-\frac{\theta N_{A}}{V} \right)\left( 1-\theta\right) \right]-k_{off}\theta$$

(3)

The initial state, at *t* = 0, is *θ* = 0. At the end of the primary binding step, at *t = t_prim_* , the amount bound will be *θ_1_N_A_*, which is to be computed by integration of equation (3).

First washing step: During the first washing step, analyte will dissociate from the surface.

$$\frac{d\theta}{dt}=-k_{off}\theta$$

(4)

The initial state, at *t* = 0, is *θ = θ_1,t = tprim_*. After time *t = t_wash_*, the amount remaining will be *θ_2,0_*, the starting condition for the next step,which is computed by integration of equation (4):

$$\theta_{2,0}=\theta_{1,t=t_{prim}}\text{exp}\left( -k_{off}t_{wash} \right)$$

(5)

Secondary binding step: The secondary binding step is described in the same way as the first, with initial secondary reagent concentration *c_sec,0_* , and with reaction rate constants *k_on,sec_* and *k_off,sec_* . The binding is to sites on the surface that have target analyte bound following the primary binding step and the first wash. Two processes are happening. First, the target analyte continues to dissociate from the surface, so the available number of binding sites decreases with time. Since the amount bound is small, the concentration resulting in solution will be small and is assumed to be zero. Second, the secondary (indicator) reagent will be binding to those primary molecules that remain on the surface. It will also be dissociating as will the target to which it is bound :

$$\frac{d\theta_{2}}{dt}=\left[ k_{on,sec}\left( c_{sec,0}-\frac{\theta_{2}N_{A}}{V} \right)\left( \theta_{2,0}-\theta_{2} \right) \right]-k_{off,sec}\theta_{2}$$

And the target is also dissociating from both sites carrying indicator and those which are not

$$\frac{d\theta_{2,0}}{dt}=-k_{off}\theta_{2,0}$$

(6)

If the secondary reagent concentration is sufficiently high, then the amount bound can be ignored in relation to the amount present in solution, in which case:

$$\frac{d\theta_{2}}{dt}=\left[ k_{on,sec}c_{sec,0}\left( \theta_{2,0}-\theta_{2} \right) \right]-k_{off,sec}\theta_{2}-k_{off}\theta_{2}$$

(6a)

The initial state, at *t* = 0, is *θ* = *θ_2,0_* . At the end of the secondary binding step, at *t = t_sec_* , the amount bound will be (*θ_3_N_A_*)_t = tsec_ , which is the initial state for the next step and which is to be computed by integration of equation (6).

Second wash step . During the second wash step, time *t_wash_* , the indicator reagent can be lost both by dissociation of the indicator-analyte complex and by dissociation of the analyte from the surface.

$$\frac{{d\theta}_{2}}{dt}=-\left( k_{off}+k_{off,sec} \right)\theta_{2}$$

(7)

The amount remaining after the wash time would be

$$\theta_{2,final}N_{A}=\left( \theta_{3}N_{A} \right)_{t=t_{sec}}exp\left( -\left( k_{off}+k_{off,sec} \right)t_{wash} \right)$$

(8)

This is the amount that is measured when the indicator is measured and is therefore the assay result.

The parameters that can be controlled to tune the assay result are the number of binding sites on the capture surface, *N_A_* , the times *t_prim_* , *t_sec_* , and *t_wash_* , and the concentration of the secondary (indicator) reagent, *c_sec,0_* .

3. *Model results*

The equations are solved numerically. For the purpose of modelling the behaviour it is reasonable to assume that the binding and dissociation rate constants for the primary and secondary steps are so similar that they can be taken to be the same. This conveniently reduces the number of variable parameters and simplifies the scaling needed for numerical solution.

Time and concentration are scaled to be dimensionless. Time is taken as the ratio to the ‘on’ time, *τ = V/(k_on_N_A_)* . and concentration to *N_A_/V* which is the ‘concentration’ of binding sites in relation to the solution volume. Denoting the scaled time by *T* and concentration by *Γ*:

Primary binding:

$$\frac{d\theta}{dT}=\left( \Gamma-\theta\right)\left( 1-\theta\right)-K_{off}\theta$$

(9)

First wash:

$$\theta_{2,0}=\theta_{1, T=T_{prim}}\text{exp}\left( -K_{off}T_{wash} \right)$$

(10)

Secondary binding:

$$\frac{d\theta_{2}}{dT}=\Gamma_{sec}\left( \theta_{2,0}-\theta_{2} \right)-K_{off}\theta_{2}$$

$$\frac{{d\theta}_{2,0}}{dT}=-K_{off}\theta_{2,0}$$

(11)

Secondary wash:

$$\theta_{2,final}={\theta_{2, T=T}}_{sec}exp\left( -2K_{off}T_{wash} \right)$$

(12)

The final result is the dependence of *θ_2,final_* on *Γ*, with the variable parameters *Γ_sec_* , *T_prim_ , T_sec_* , and

*T_wash_*

Figure 1 shows the time-variation of coverage of captured analyte in the primary binding step. At longer time, the signal saturates: this is the regime for an equilibrium assay. At sufficiently short time, the increase of coverage is linear with time and at fixed time is linear in concentration. This is the regime for a kinetically-controlled assay. In this regime, the precision of the assay result will be determined by the precision with which time zero is known – hence the need for rapid and efficient mixing of the sample solution with the capture surface – and by the precision of timing the end of the primary incubation step- hence precision and speed in flushing the sample solution away from the surface, fast introduction of the wash and rapid and efficient mixing in the wash step. Figure 2 shows the time variation of the coverage of the secondary indicator, with varying secondary reagent concentration, for a particular target analyte concentration, primary incubation time, washing time and dissociation rate constant. The dissociation of the indicator off the capture surface can be an important issue and there is an interaction between the effects of secondary incubation time and secondary reagent concentration. For a reliable assay system, ideally the result should be independent of the secondary reagent concentration. Figure 2 shows that this can be achieved if the secondary reagent concentration is high enough and the incubation time is long enough. Longer incubation leads to loss of signal but higher secondary reagent concentration could lead to issues of non-specific binding of this reagent which would then lead to erroneously high assay results. The effect of dissociation is to impose strong requirements on the precision of timing for both the wash and secondary incubation steps.

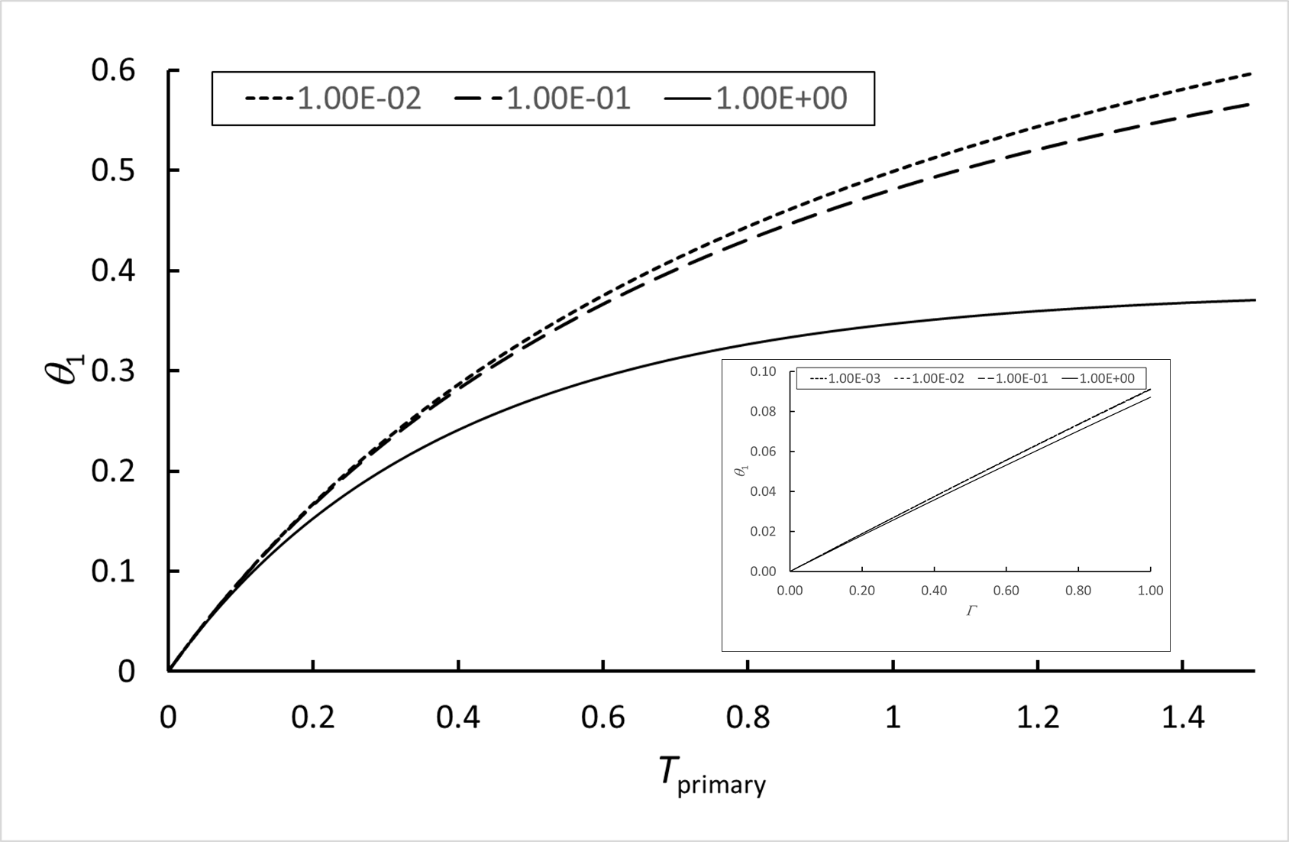

*Figure 1* Variation of surface coverage of analyte captured during the primary binding step, *θ_1_* , with time, *T = tk_on_N_A_/V* Inset: variation of *θ_1_* at *T_primary_* = 0.1 with scaled analyte concentration, *Γ =c_0_V / N_A_*. Parameter: “off” rate, *K_off_ = k_0ff_V/k_on_N_A_* .

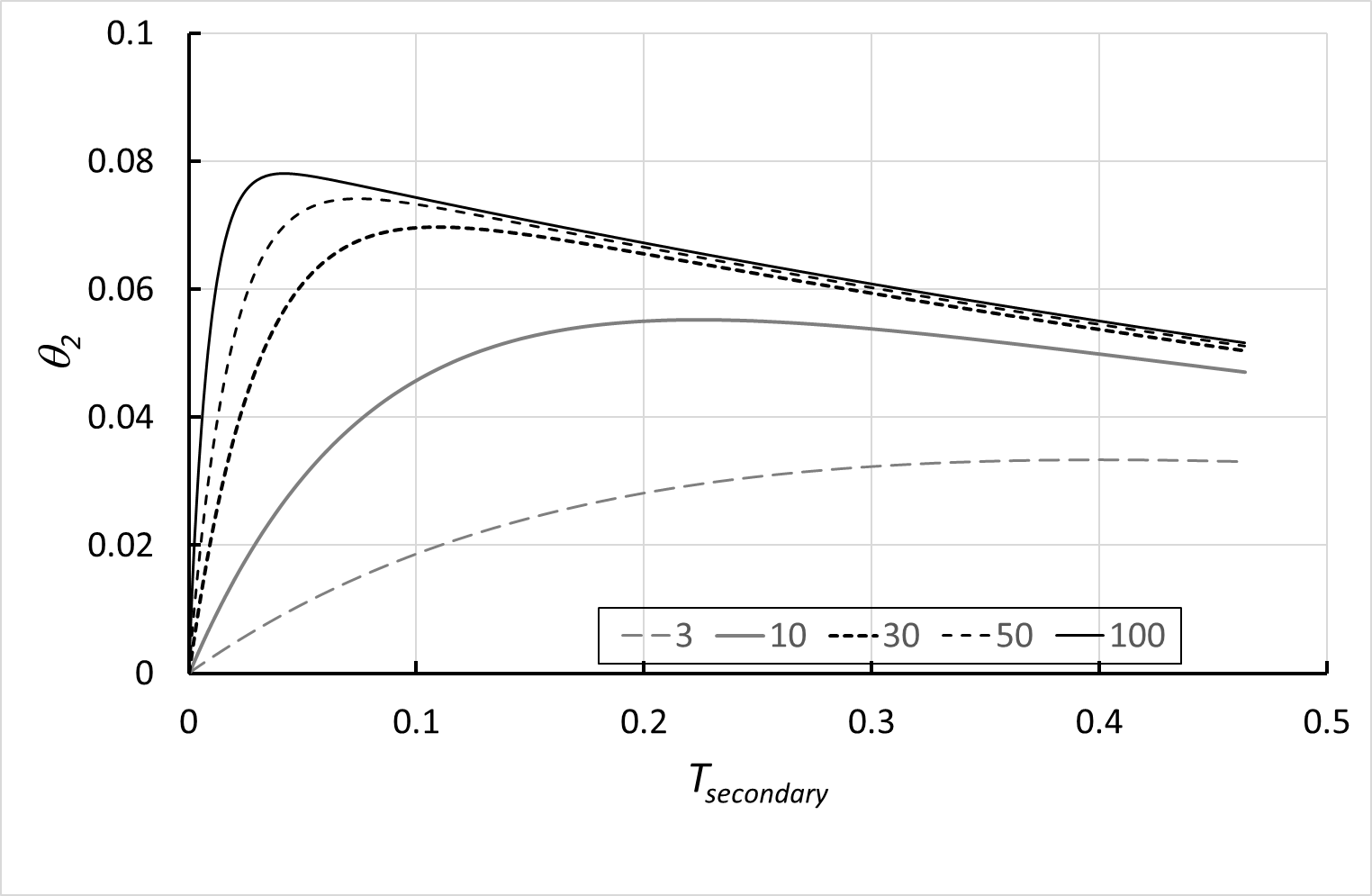

Figure 2*.* Time variation of the surface coverage of the secondary (indicator) reagent, *θ*_2_ , with different scaled secondary reagent concentration (legend). Scaled analyte concentration, *Γ* = 1; primary incubation time, *T_primary_* = 0.1, scaled ‘off’ rate, *K_off_* = 1 . The decay at longer time is a consequence of the dissociation from the surface of the bound target analyte and of the secondary reagent from the bound target.

The principal determinant of the assay sensitivity and dynamic range is the amount of analyte captured in the primary incubation step, *θ*_1_ . If *K_off_* is small, then equation (9) has solution:

$$\theta_{1}=\frac{\Gamma\left( 1-exp(-T(1-\Gamma) \right)}{\left( 1-\Gamma exp(-T(1-\Gamma) \right)} \Gamma\neq1$$

(13)

So if *T*(1-*Γ*) is small : *θ*_1_ = *Γ T* .

And can be approximated to second order:

$$\theta_{1}\approx\frac{\Gamma T}{\frac{1}{1-({T\left( 1-\Gamma\right)}/2)}+\Gamma T}\approx\frac{\Gamma T}{1+\Gamma T}$$

(14)

That is, the assay will noticeably deviate from linearity when *ΓT* is large enough, say *ΓT* ~ 0.1 . Figure 3 shows a numerical solution example compared with equations (13) and (14). Equation (14) is a useful approximation for fitting the assay data and thereby extending the dynamic range.

The assay can be tuned for sensitivity and dynamic range by altering the scaling factors for time, *τ*=*V/(k_on_N_A_)* and concentration, *N_A_/V* , which is conveniently done by altering the sample volume, *V*, and loading of the capture species on the capture bead, *N_A_*. .

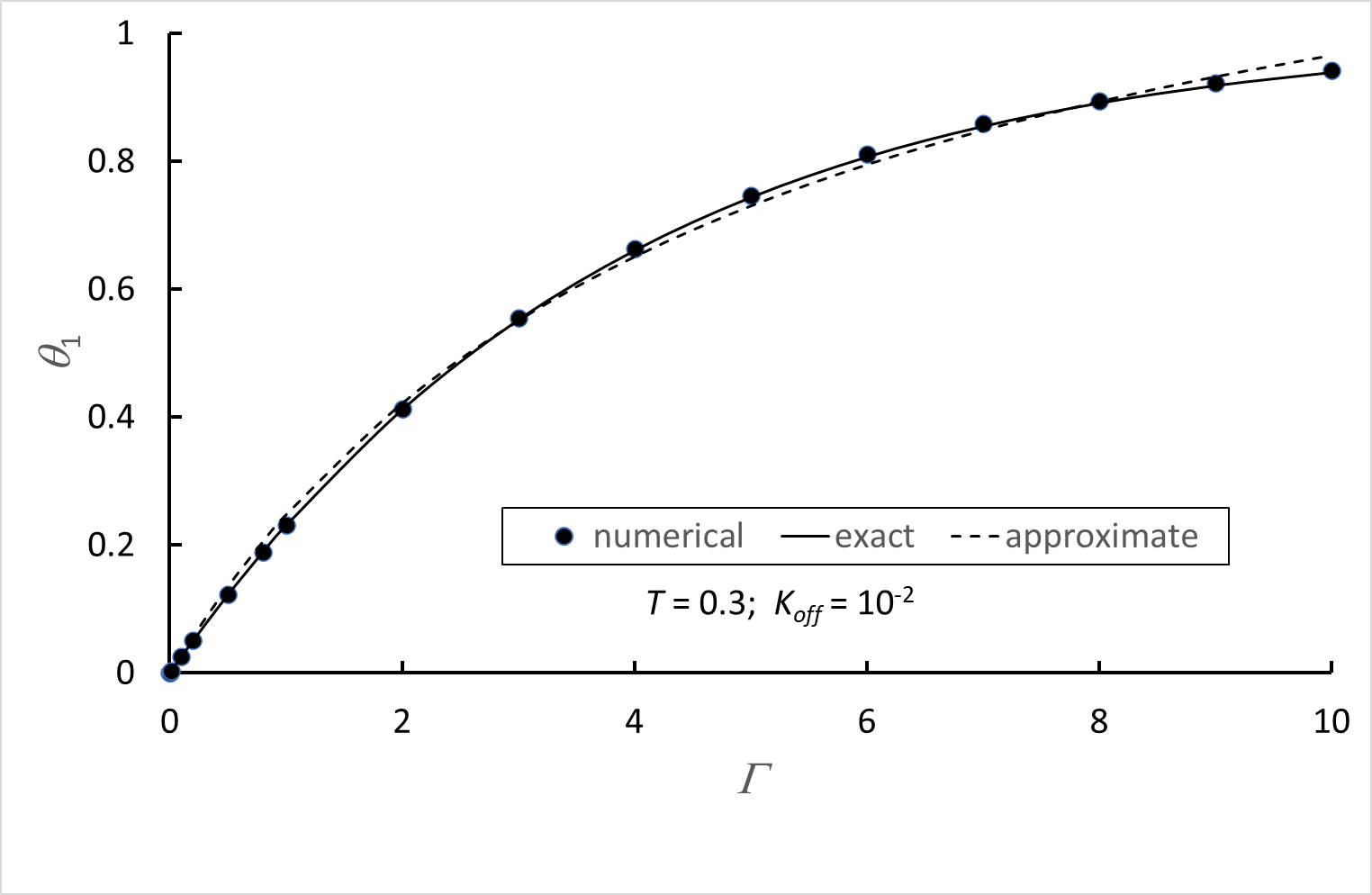

Figure 3. Numerical solution of equation (9) compared with equations (13) (labelled ‘exact’) and (14) (labelled ‘approximate’).

*Signal generation*

The assay is an implementation of a standard Enzyme-Linked Immunosorbent Assay (ELISA) with colorimetric output. There are 5 steps:

a) Capture of the antibody to be measured onto a surface carrying the antigen specific for the antibody. For the SARS-CoV-2 assay, the antigen is the Receptor Binding Domain (RBD) protein. (SARS-CoV-2 (2019-nCoV) Spike RBD-His Recombinant Protein, Biotinylated -Sino Biologicals 40592-V08H-B)
b) Washing off all unbound material
c) Incubation of the capture surface with horseradish peroxidase (HRP) – labelled anti-human IgG (Goat anti-Human IgG Fc Secondary Antibody HRP - Thermofisher - A18817)
d) Washing off unbound material
e) Incubation with a solution of tetramethyl benzidine and hydrogen peroxide (TMB) and measurement of the variation over time of the blue colour that is developed.

The control antibody is SARS-CoV-2 Spike Protein (S-ECD/RBD) Monoclonal Antibody (bcb01) - Thermofisher - MA5-35948

The key to the assay accuracy is rapid and complete mixing and rapid timing. The assay result is controlled by the kinetics of binding in steps a) and c) and the speed of washing in steps b) and d). With these fixed and under control, the development of colour in step e), determined by the amount of HRP bound onto the surface, can reliably be related back to the concentration of antibody present in the sample solution. The objective is to determine, from the time dependence of the result, the rate constant for colour development relative to that for a control sample of known concentration.

a) Expected time-dependence of assay signal

The assumption is that the concentration of hydrogen peroxide is sufficiently high that it does not influence the rate of conversion of colourless TMB to its blue-coloured product. Letting *c_TMB1_* denote the concentration of the blue product (concentration at the start of the incubation is zero and at the end is equal to the concentration of colourless TMB added, denoted *c_TMB_* ), the conversion is assumed to be a first-order chemical reaction with rate constant proportional to the concentration of HRP that is present.

$$c_{TMB1}=c_{TMB}\left( 1-exp\left( -kt \right) \right)$$

(15)

Where *t* is the elapsed time since addition of TMB and *k* is the rate constant. Letting *k_control_* and *c_control_* denote respectively the rate constant determined for the control sample and the concentration of the control sample, then the concentration of the unknown sample, *c_unknown_* , with measured rate constant *k_unknown_* is:

$$c_{unknown}=c_{control}\frac{k_{unknown}}{k_{control}}$$

(16)

Concentration is measured by measuring the intensity transmitted through the solution of light of wavelength matching the wavelength of maximum absorption of the blue product. The relationship of light intensity to concentration would be given by Beer’s law (*I* denotes measured transmitted light intensity, *I_0_* the source intensity, *L* the length of the light path through the solution and *ε* the absorption coefficient of the blue product:

$$\frac{I}{I_{0}}=exp(-\varepsilon Lc)$$

(17)

However, if the light path,*L*, is sufficiently short that the amount of light absorbed is small then a simplifying approximation can be made:

$$\frac{I}{I_{0}}\approx1- \varepsilon Lc$$

(18)

In that case, the variation of light intensity with time would be:

$$\frac{I_{0}-I}{I_{0}-I_{\infty}}=(1-exp\left( -kt \right)$$

(19)

Where *I_0_* and $I_{\infty}$ here denote the light intensity measured at *t* = 0 (immediately after mixing) and $I_{\infty}$ is the light intensity at time sufficiently long to convert all the added TMB to its blue product.

b) Confirmation of expected time dependence of signal

Figure 3 shows an assay result for a concentration at the upper end of the required measurement range, where the assay signal – output of a photodiode measuring transmitted light intensity – covers almost the full range. The lines show non-linear least squares fit of the light intensity vs time assuming a first-order increase with time of concentration of the blue product (eq 15) and either Beer’s Law (eq 17) or the linear approximation (eq 18) for the dependence of light intensity on concentration. The fit is reasonable. The rate constant for generation of the blue colour relates directly to the amount of bound secondary indicator (HRP), *θ_2,final_N_A_* which relates to the amount of target captured in the primary incubation step and hence to the target analyte concentration.

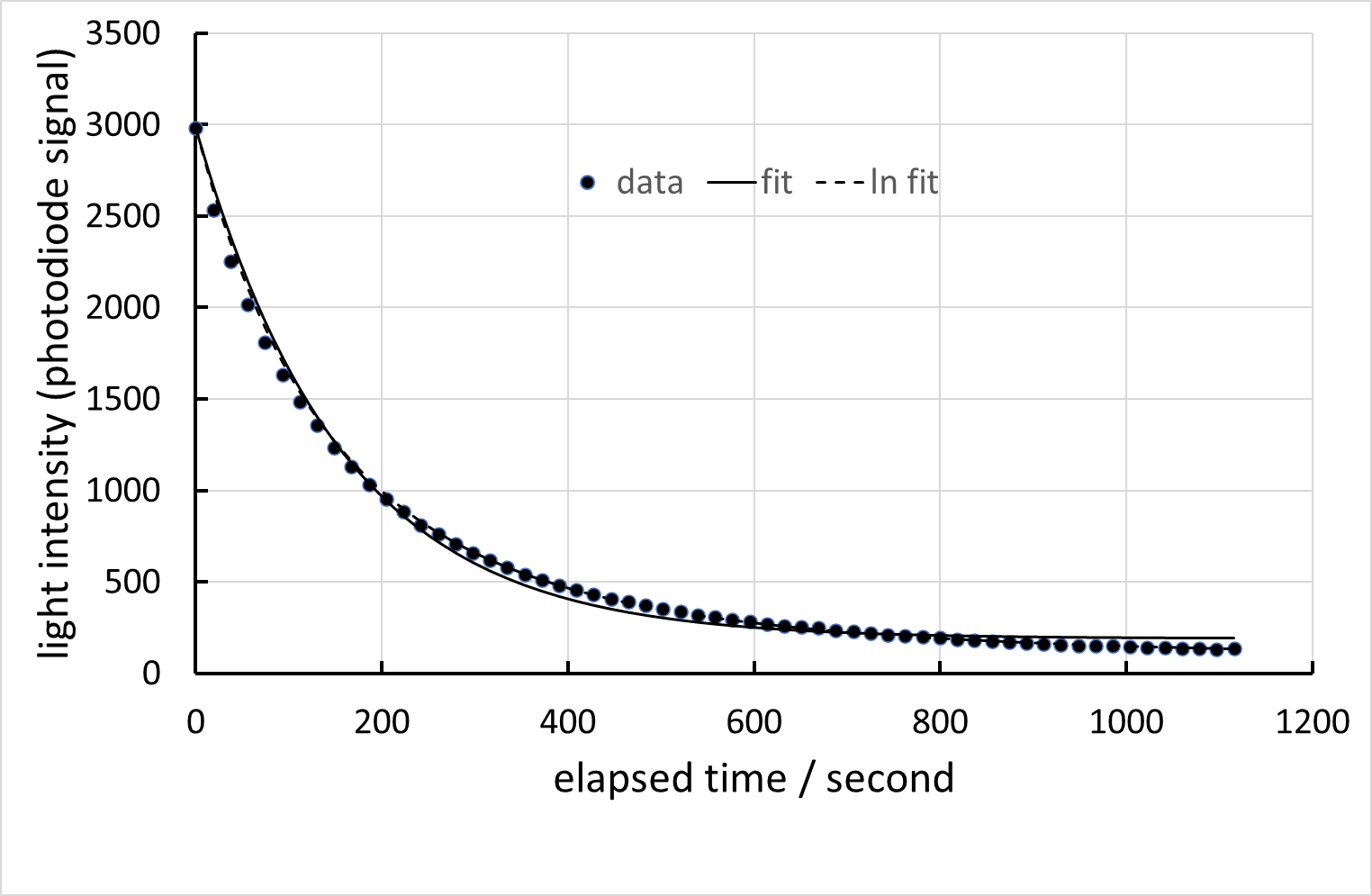

*Figure 3 Measured light intensity vs elapsed time from addition of TMB (points) and fit to expected first-order variation assuming Beer’s Law (dashed line) and the linear approximation (full line). Analyte concentration 10μg / mL*

c) Confirmation of expected dependence of rate constant for signal generation on target concentration and demonstration of wide dynamic range.

Figure 4(a) shows assay signal development with different concentration of target in the diluted solution. The rate constant for signal development increases smoothly with increasing target concentration. Figure 4(b) illustrates the performance of a fitting algorithm designed to progress smoothly from the linear variation at low analyte concentration to the exponential variation at high analyte concentration (equation 19). The desired result is the initial slope relative to that for a control on the same disc (equation 16). Figure 5 shows the variation of derived rate constant with target analyte concentration. The variation is consistent with the approximate solution, equation (14). Saturation of the signal is caused by saturation of the capture surface (*θ*_1_ 🡪 1). The dynamic range is over a scale of about 200 times in concentration. The linear range is over a range of approximately 20 times in concentration. As noted above, the assay range can be adjusted primarily by altering the loading of the capture antigen onto the capture surface. The difference between duplicate measurements is 10% of the measurement across the assay range and would be largely determined by the timing accuracy. The assay is sufficiently sensitive that the sample can be heavily diluted with buffer, which is helpful in normalising samples as variable as blood.

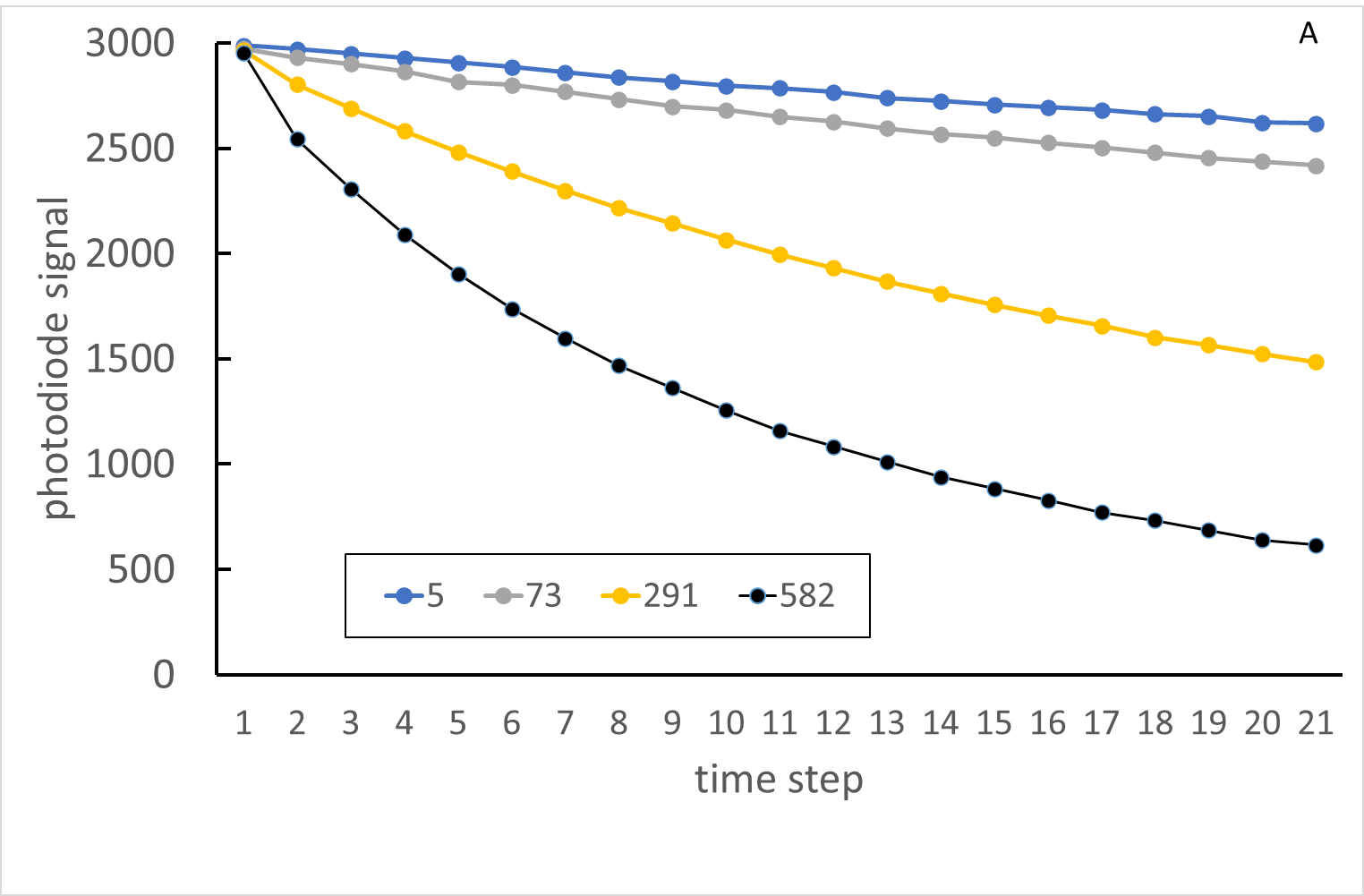

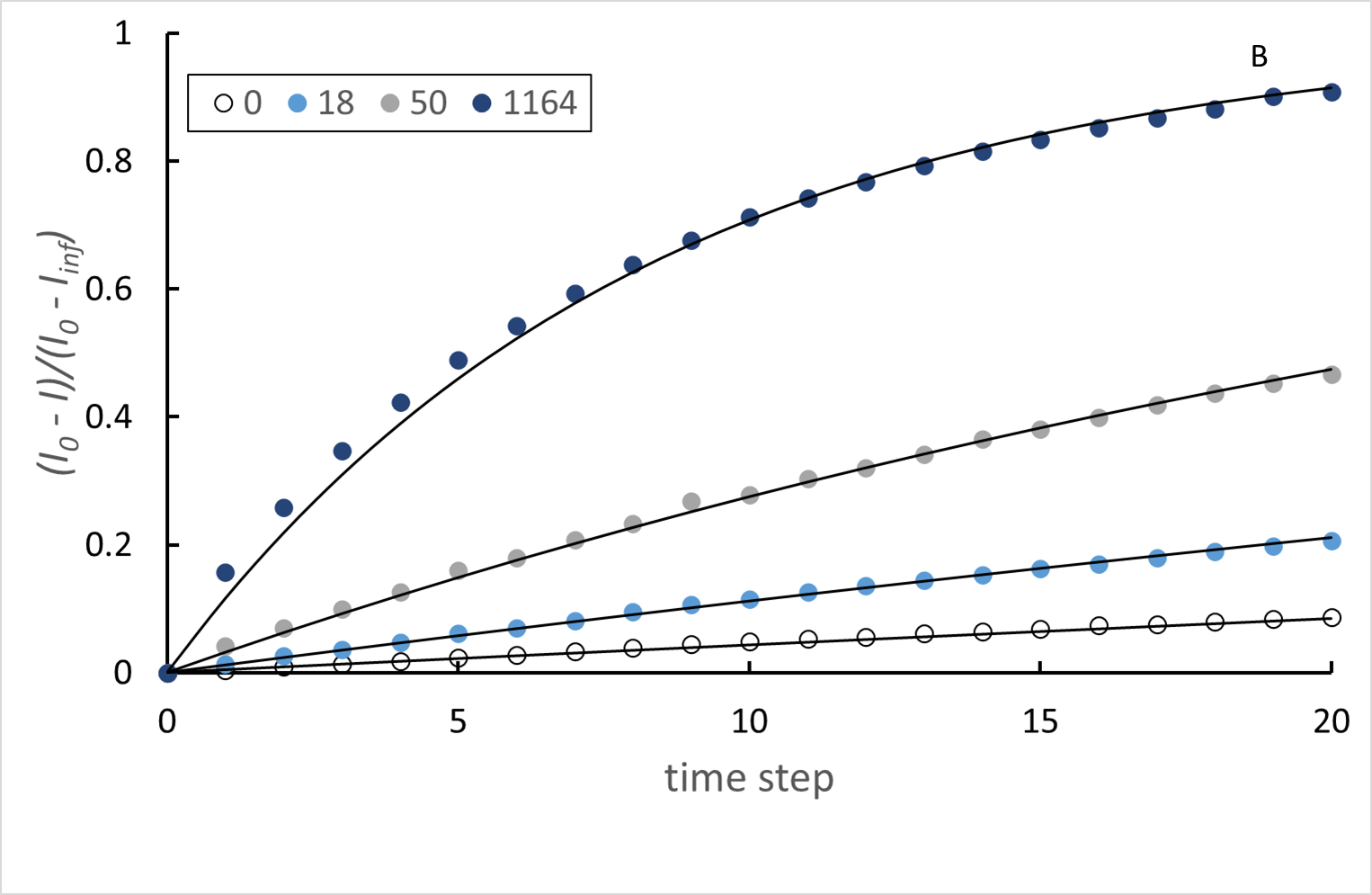

*Figure 4*. *A: Variation of colour development with change of target analyte concentration. The rate constant is extracted as the assay signal. Legend: diluted target concentration, ng / mL. The time step is 18 s. B: Variation according to equation (19) Lines are fitted with an algorithm designed to move smoothly from the linear regime at low analyte concentration to the exponential regime at high concentration*

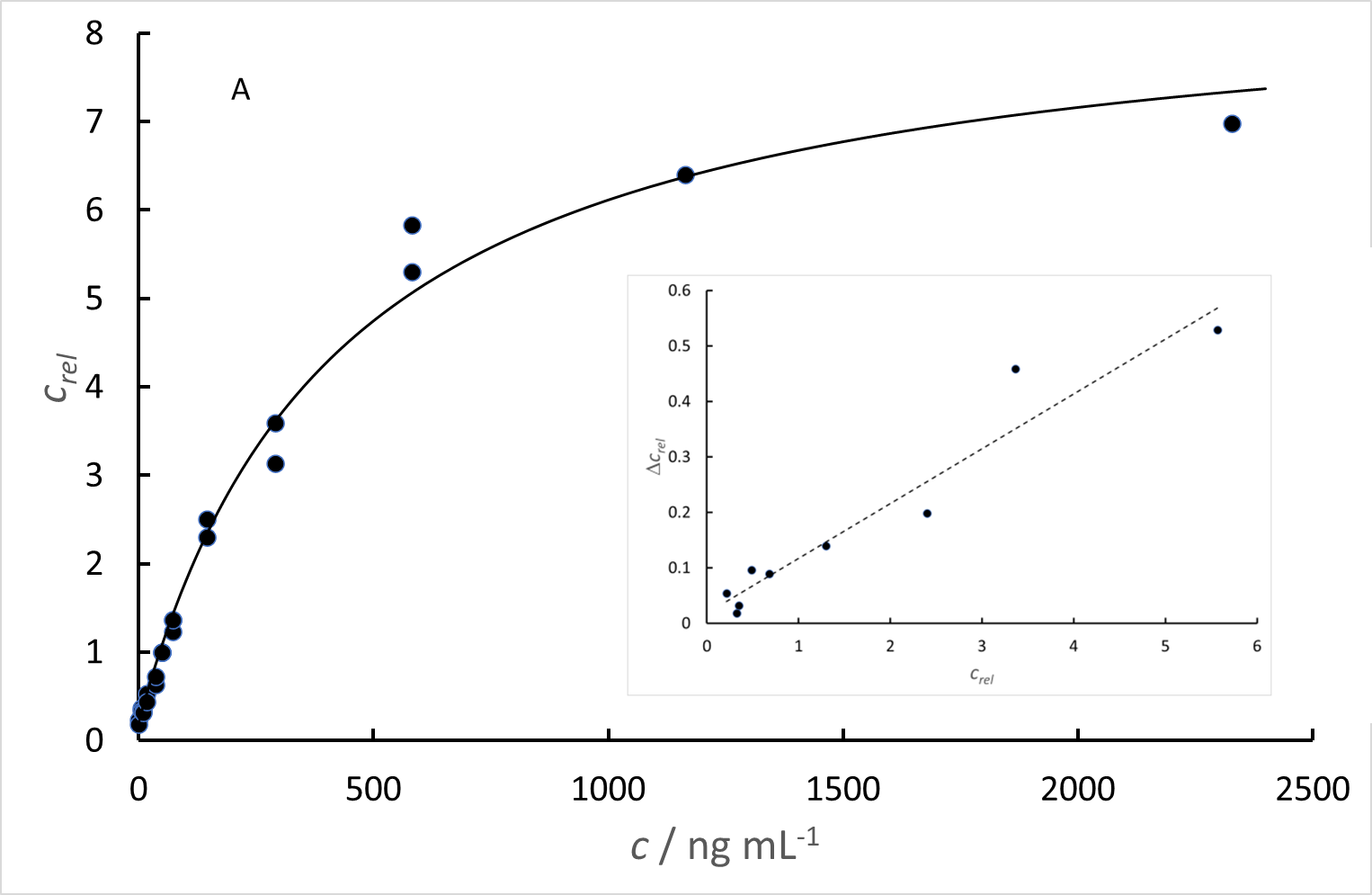

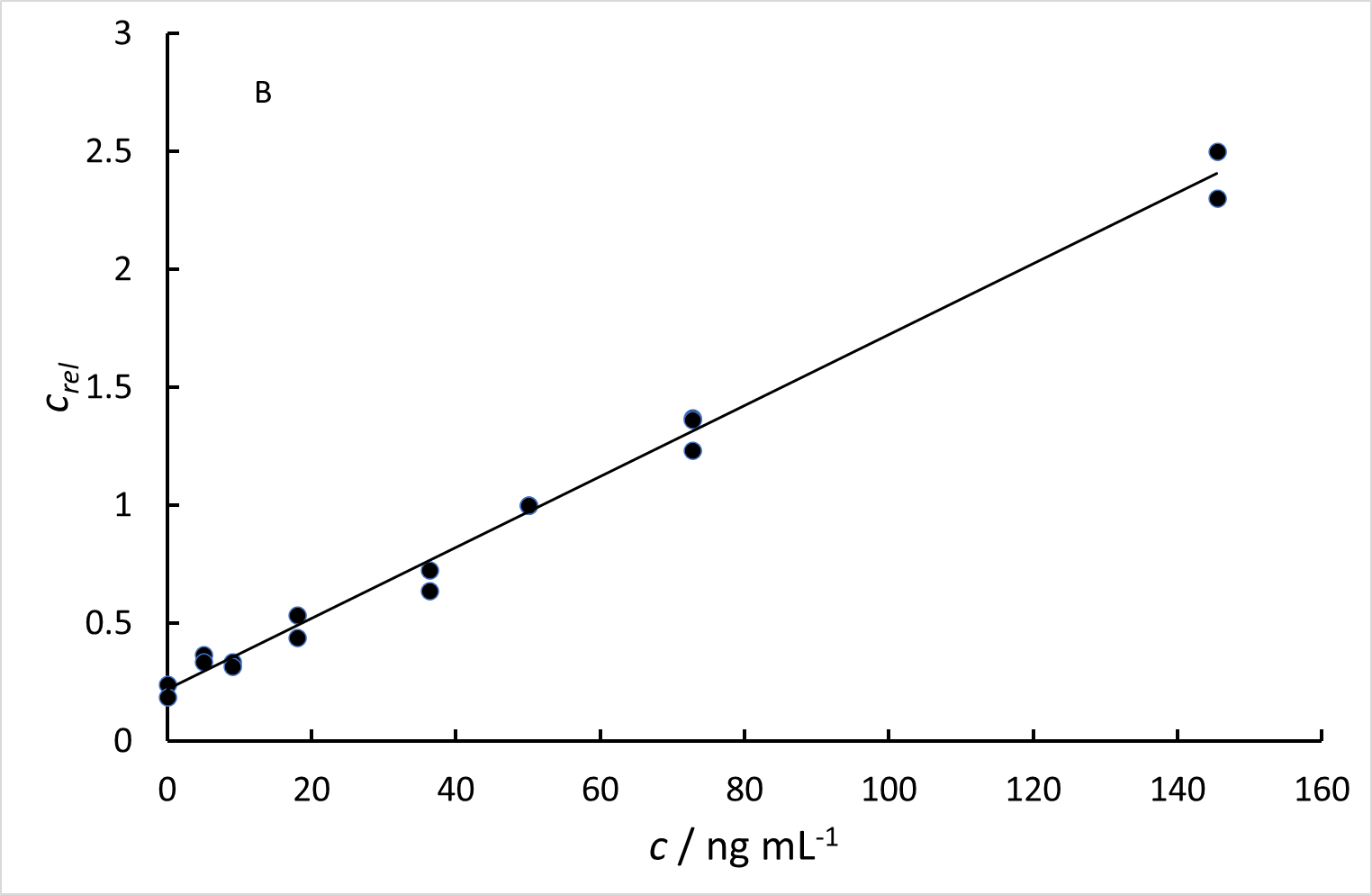

*Figure 5. A. Variation of relative concentration c_rel_ (determined as rate constant for colour development relative to the control - 50 ng / mL). The line is a fit to the approximate solution of eq (14). Inset: Difference between duplicate measurements relative to the control, Δc_rel_ as a function of the measured concentration relative to the control, c_rel_ ; the assay shows a coefficient of variation of 10% that is essentially constant across the assay range. B: linear range at low target analyte concentration; concentration relative to the control, c_rel_, against sample concentration, c*.

Supplementary Appendix 3: Assay validation with WHO standards.

Panel 20/770 from the National Institute for Biological Standards and Control, UK, (NIBSC) comprises a set of plasma samples from convalescent Covid-19 patients as well as a set of pre-Covid negative samples. With the panel is provided a set of analytical results performed on a variety of different medical laboratory assay platforms. Because all the platforms have a different concentration scale, for the comparisons below, these results have been scaled to the arithmetic for the positive samples in the set, determined for each platform. Not all of the assays are directed against the same antigen: notably the assays from Abbot and Roche used for evaluation of the panel. For comparison, assays directed against the S1 protein or RBD are used. The in-house assay from Public Health England, Colindale Laboratory, is taken as the comparator. Figure 1 shows the correlation of the various assays with the PHE Colindale result. The Euroimmun and Ortho assays correlate well with PHE Colindale. The Siemens assay against the RBD correlates less well and is restricted by its limited dynamic range. Abbott and Roche assays, being directed at different antigens, do not correlate well with PHE Colindale.

Figure 2 shows the correlation of the Orbis assay result with PHE, including a direct comparison with the Siemens result. On the Orbis assay, the negative plasma samples showed the same non-specific binding offset observed for the clinical study negatives, with closely similar mean and standard deviation. The mean offset was therefore subtracted from the assay result. There are two error contributions: the variability in non-specific binding, and the constant ~10% replication error (see Supplementary Appendix 3). Orbis and Siemens show similar correlation, with a correlation slope close to unity.

One question is whether the difference in results for the Orbis assay compared to the other assays presented here relates to differences in assay design, in response to samples containing a range of antibodies of different affinity for the chosen target. Plate-based assays are usually incubated to equilibrium (at least 1 hr) and therefore develop results reflecting the antibody affinity. In contrast, the Orbis assay is designed for speed to result, with incubation time 5 minutes. The result is determined by the ‘on’ rate for the antibody, which may not be the same for antibodies of the same affinity. To assess this possibility, the difference between Orbis or Siemens result and the PHE Colindale result is plotted against sample number, to see whether any sample might be a significant outlier. Figure 3 shows the result. The 95% confidence interval (2 times standard deviation) for the Siemens result is taken as an estimate of inter-sample variability caused by the use of RBD rather than S1 as the antigen. Some Orbis results fall outside this variability band, though not by more than the estimated measurement error so person-to-person variability in antibody composition does not seem to be a major issue.

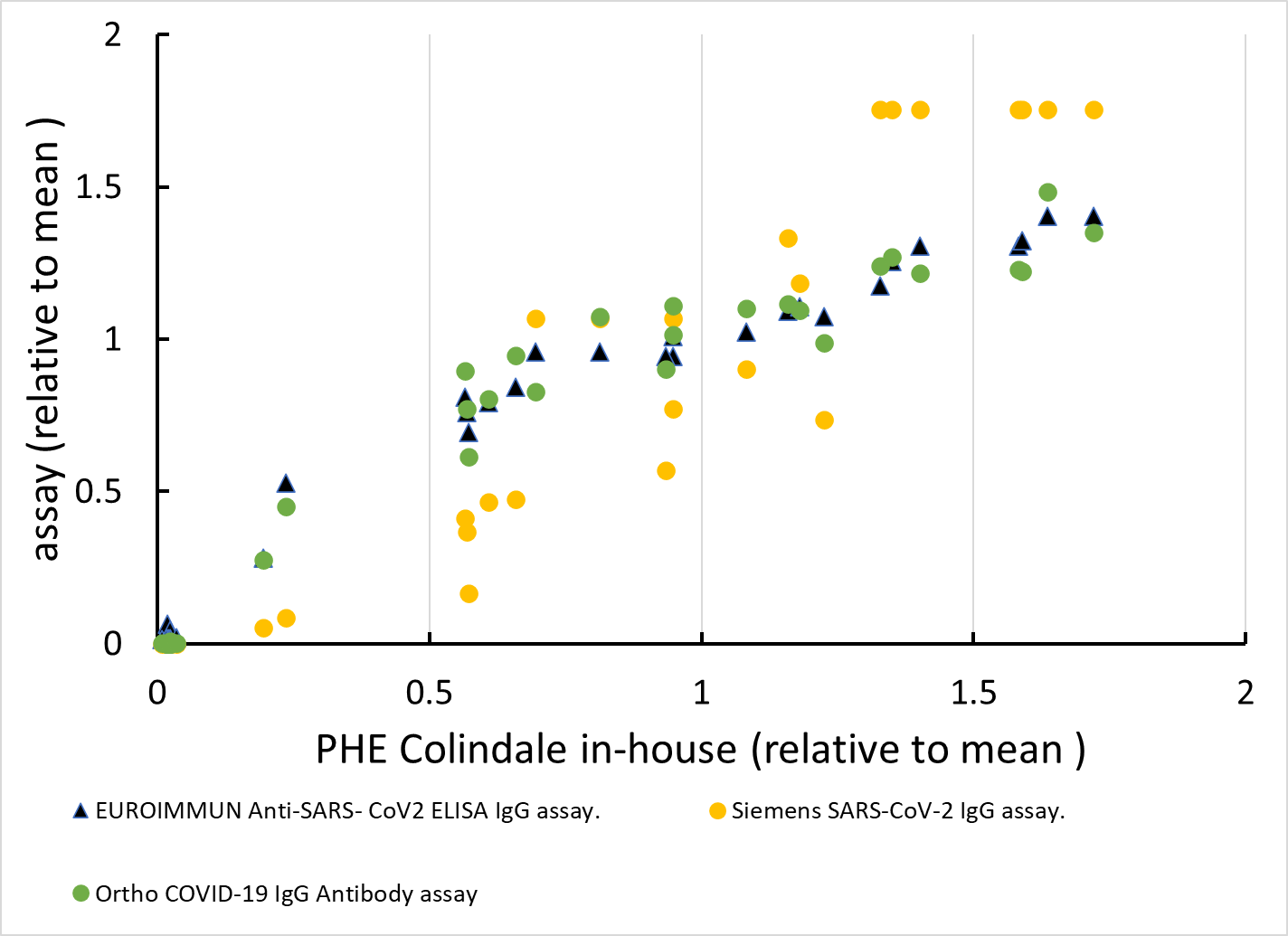

*Figure 1. Correlation of results from different medical laboratory immunoassay platforms for S1 IgG (EuroImmun, Ortho) or RBD IgG (Siemens) for the NIBSC 20/B770 panel, taking the PHE Colindale assay as the comparator; data from the 20/B770 package insert.*

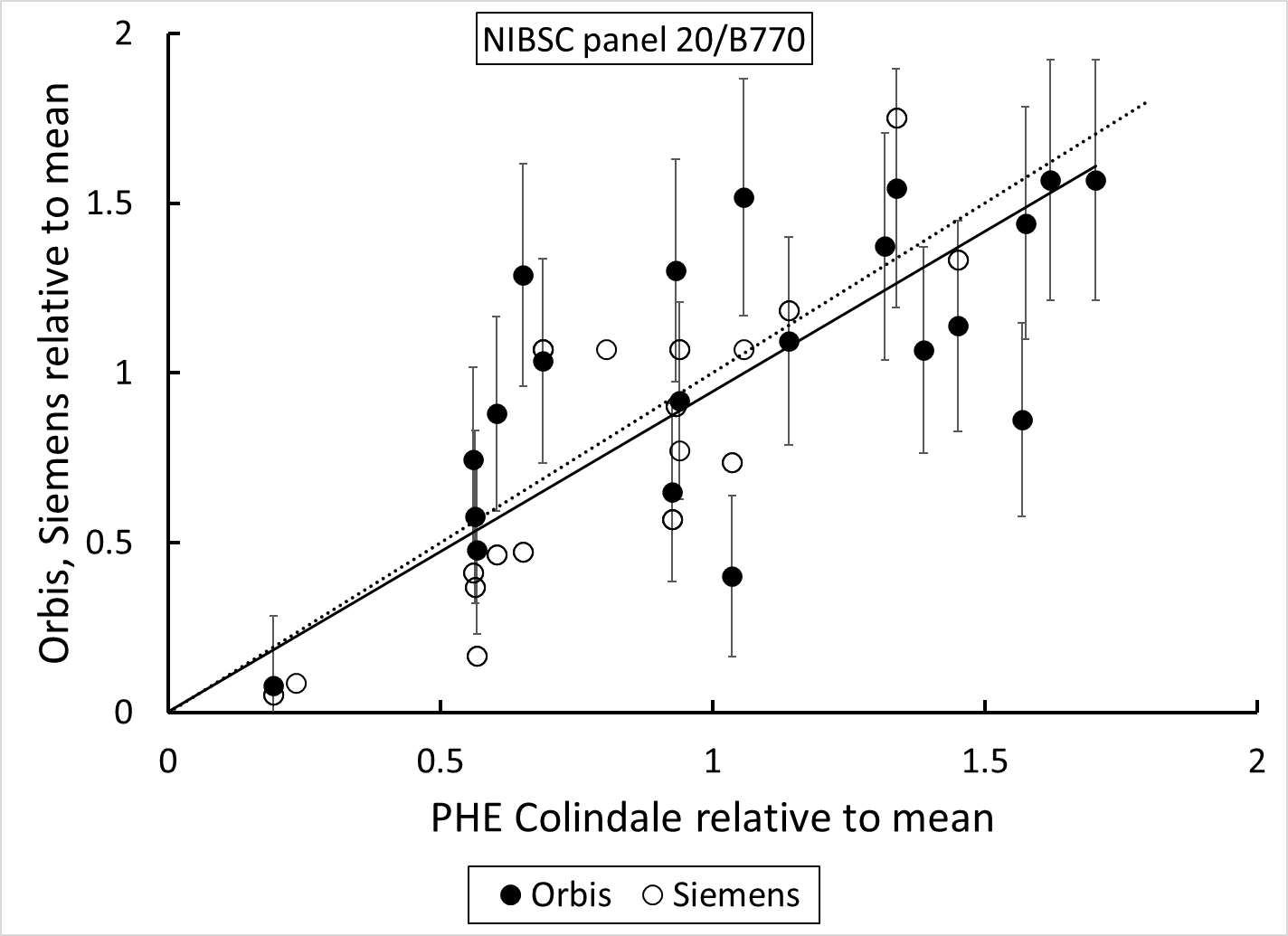

*Figure 2 Correlation of Orbis assay result (sample / control, non-specific binding offset corrected) with the result for PHE Colindale for panel 20/B770; Comparison with Siemens anti-RBD result from the 20/B770 package insert, where reslts showing greater than the assay dynaic range have been excluded. The lines are: dotted – 1:1 correlation; solid- Orbis. Error bars are 95% confidence limits assessed from the replication error and non-specific binding error*

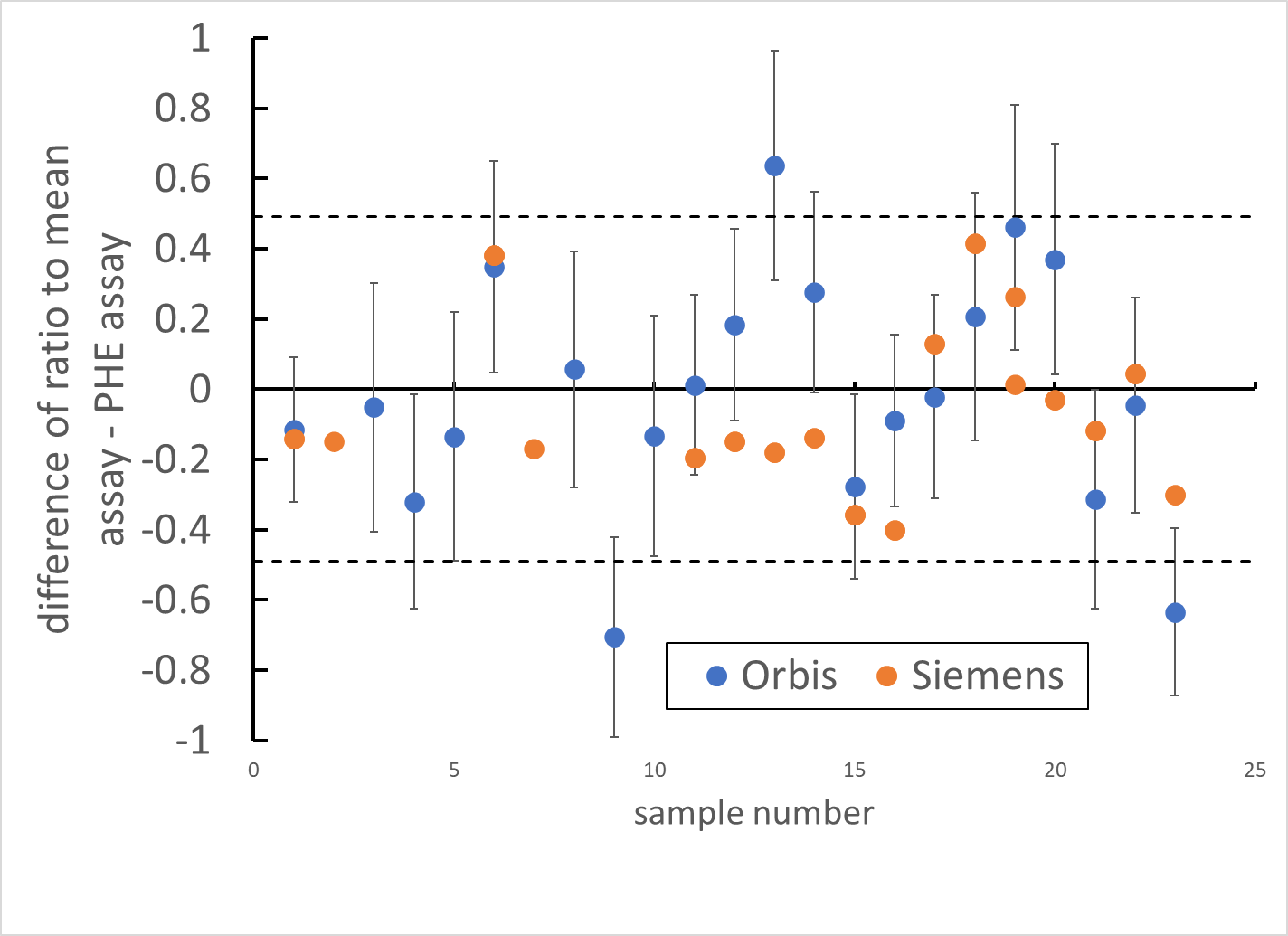

*Figure 3. Comparison of Orbis and Siemens assay result by sample: difference between assay result and PHE Colindale assay result, where results are expressed as the the ratio to the mean positive result. Dashed lines are 2 times the standard deviation of differences for the Siemens result.*

In order to express the Orbis assay results on a recognised comparison scale, the standard reagents NIBSC 20/162 and the panel 20/150 were measured. Figure 4 shows the result.

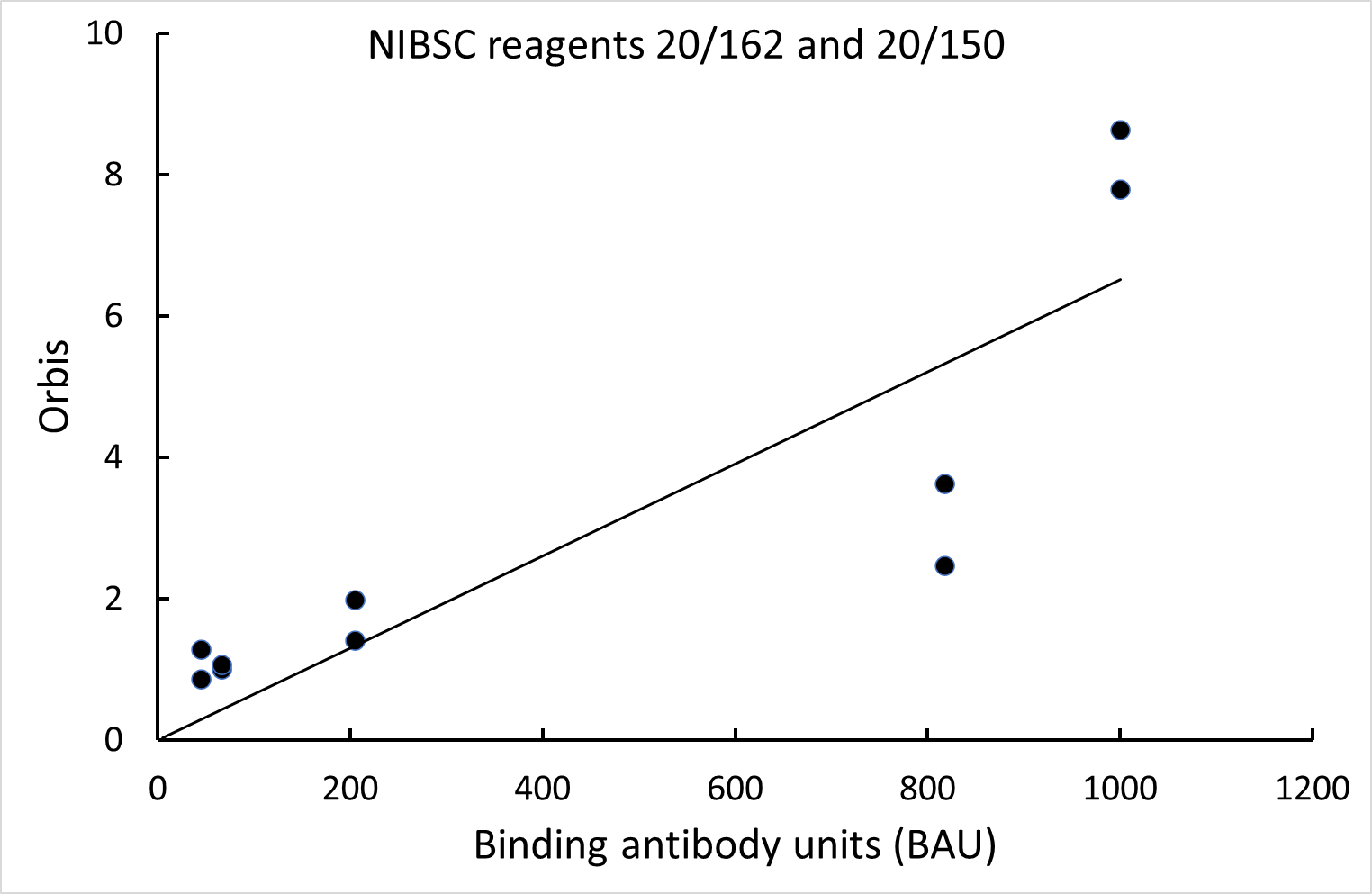

*Figure 4. Orbis assay (non-specific binding offset corrected) results for WHO standard samples*

The NIBSC gives concentration of anti-receptor binding domain IgG in ‘binding antibody unit’ (BAU) derived for each sample as the median (geometric mean) of the results from a number of different assay methods. The samples, being derived from convalescent plasma, would contain antibodies with a range of different binding affinity that may react differently in different assays dependent on the details of the assay design. The range of antibodies present would be different in each sample. Exact agreement is not therefore to be expected.
